# Computational Mechanisms Underlying Multi-Step Planning Deficits in Methamphetamine Use Disorder

**DOI:** 10.1101/2024.06.27.24309581

**Authors:** Claire A. Lavalley, Marishka M. Mehta, Samuel Taylor, Anne E. Chuning, Jennifer L. Stewart, Quentin J. M. Huys, Sahib S. Khalsa, Martin P. Paulus, Ryan Smith

**Author notes:** Corresponding author (Ryan Smith). Laureate Institute for Brain Research. 6655 S. Yale Ave. Tulsa, Oklahoma, 74136, United States.

## Abstract

Current theories suggest individuals with methamphetamine use disorder (iMUDs) have difficulty considering long-term outcomes in decision-making, which could contribute to risk of relapse. Aversive interoceptive states (e.g., stress, withdrawal) are also known to increase this risk. The present study analyzed computational mechanisms of planning in iMUDs, and examined the potential impact of an aversive interoceptive state induction. A group of 40 iMUDs and 49 healthy participants completed two runs of a multi-step planning task, with and without an anxiogenic breathing resistance manipulation. Computational modeling revealed that iMUDs had selective difficulty identifying the best overall plan when this required enduring negative short-term outcomes – a mechanism referred to as aversive pruning. Increases in reported craving before and after the induction also predicted greater aversive pruning in iMUDs. These results highlight a novel mechanism that could promote poor choice in recovering iMUDs and create vulnerability to relapse.

## Introduction

Methamphetamine dependence has devastating physical and psychological consequences (1) and rates of overdose and mortality have continued to increase in recent years (2, 3). The growing prevalence of methamphetamine use disorder, poor treatment outcomes, and high rates of relapse all necessitate a better understanding of its etiology and maintenance factors (4, 5). While various risk factors have been identified (e.g., dependence on more than one drug, family history (6, 7)), mechanisms promoting continued use despite negative consequences remain inadequately understood.

Maladaptive decision-making processes represent one mechanism expected to contribute to continued substance use and relapse. Current research examining decision-making in meth/amphetamine users has found greater impulsive behavior (8, 9), impaired inhibitory control, and increased risk-taking compared to healthy participants (10, 11). Impulsivity in methamphetamine users has also been shown to positively predict craving severity (12). Heightened impulsivity and reduced inhibition may reflect a lack of consideration for, or even an impaired ability to consider, potential consequences before choices are made. In support of this idea, methamphetamine users demonstrate heightened delay-discounting compared to non-users (13, 14). While discounting has been examined in various substance use disorders, the related trait of *cognitive reflectiveness* (15–17) has received little attention in this population and may offer further explanation for impaired decision-making (i.e., by failing to consider distal outcomes). The substance use literature has instead tended to focus on reward processing. Along these lines, a recent neuroimaging study in individuals with amphetamine use disorder showed greater anticipatory reward processing (right amygdala activation) before large-win trials during a Monetary Incentive Delay task compared to non-users (18). This finding adds to prior literature suggesting that decision-making impairments in amphetamine use disorders may be affected by impaired reward-processing that is biased toward large, immediate reward.

Computational modeling of decision-making, which allows one to mathematically formalize cognitive processes, offers a quantitative approach for assessing behavioral tendencies and has become frequently used in substance use research in particular (for a recent review, see Smith, Taylor (19)). One computational framework, known as reinforcement learning (RL; (20)), focuses on learning from rewards and punishments to guide decision-making. Of most relevance here, there are two broad classes of RL algorithms that attempt to explain decision-making: *model-free* (MF) algorithms, which model decision-making through trial-and-error action value learning based on observed outcomes (i.e., assuming no explicit future expectations), and *model-based* (MB) algorithms, where decisions are instead made according to an individual’s internal model of the environment (i.e., which explicitly incorporates rewards expected in the future depending on choice of action). These two algorithm classes highlight the tradeoff between maximizing performance and adaptability (MB algorithms) vs. computational efficiency (MF algorithms).

To date, there has been ample discussion of possible mechanisms underlying substance abuse and the best way to model associated cognitive processes (detailed in multiple recent reviews (19, 21)). MF explanations have received more attention in experimental studies to date, and often attribute continued substance use to learned behavior (i.e., habits) based on observations of positive outcomes following drug use and negative outcomes associated with withdrawal. On the other hand, a more MB approach would explain continued use by assuming that affected individuals may overweight the expected value of a substance and underweight the expected negative consequences (22). Investigations focused on distinguishing these processes in SUDs have found evidence suggesting a shift in reliance from MB to MF decision-making (23, 24); yet, the mechanisms behind this shift, and those underpinning MB deficits, remain largely unknown. Multi-step planning, one paradigmatic example of an MB cognitive process, has not received sufficient attention in empirical studies of SUDs to date and may be crucial for understanding these impairments.

Another factor widely understood to influence decision-making and relapse is avoidance of negative affect – and avoidance of the aversive interoceptive states linked to stress and withdrawal in particular (25–29). The acute effects of methamphetamine use also have well-described sympathomimetic effects on interoceptive states (e.g., cardiovascular and respiratory tone). Planning, and potentially discounting future action outcomes, could thus be affected by the initial discomfort expected under certain options or may be preferentially impacted during such heightened negative states. For example, some decisions may not be given ample consideration due to unpleasant short-term effects (e.g., expected withdrawal states), even if longer-term outcomes would be ideal (e.g., recovery). This notion of difficulty considering possible action sequences during planning due to unpleasant short-term outcomes has been referred to as *aversive pruning* (i.e., based on the metaphor of a decision tree) and appears to be a reflexive, Pavlovian response (30). However, potential moderators of this mechanism, such as the aversive interoceptive states discussed above, are currently unknown. The degree to which aversive pruning is relevant to those with SUDs in comparison to other potentially explanatory mechanisms – such as reduced reward sensitivity or overall planning horizon (i.e., the number of steps into the future one considers) – has also not been thoroughly investigated. A better understanding of the role of these planning mechanisms in SUDs, and their potential moderators, may therefore be crucial for a full characterization of MB deficits and how they might be targeted in treatment.

In the current study, multi-step planning in individuals with methamphetamine use disorder (iMUDs) and healthy comparisons (HCs) was assessed during an anxiogenic interoceptive perturbation protocol involving inspiratory breathing resistance. We used computational modeling to assess behavior on a previously validated planning task and compared computational metrics of behavior (i.e., aversive pruning, planning horizon, and reward sensitivity) between task runs under conditions with and without the breathing perturbation, allowing assessment of the effect of interoceptive/somatic state anxiety. Computational measures were also examined in relation to severity of drug-related consequences, withdrawal, and craving. Our primary aims were to: 1) evaluate whether the aversive state induction was an effective moderator of computational planning mechanisms, 2) differentiate evidence for competing hypotheses regarding whether maladaptive planning in iMUDs is better accounted for by amplified aversive pruning, shorter planning horizon, or reduced reward sensitivity (or some combination of these), and 3) evaluate whether differences in computational planning mechanisms may be explained by trait differences in cognitive reflectiveness and potentially predict symptom severity.

## RESULTS

### Breathing-based aversive state induction was effective at increasing anxiety

Participants (iMUDs: *N*=40; HCs: *N*=49) first tested the breathing resistance mask used for anxiety induction (see **Fig. 1A**) through exposure to 6 increasing levels of resistance (i.e., 0, 10, 20, 40, 60, and 80 cmH2O/L/sec) for 60 seconds each. They then provided anxiety ratings immediately after each exposure on a scale of 0=*no anxiety* to 10=*maximum possible anxiety one could tolerate*.

**Fig. 1.**
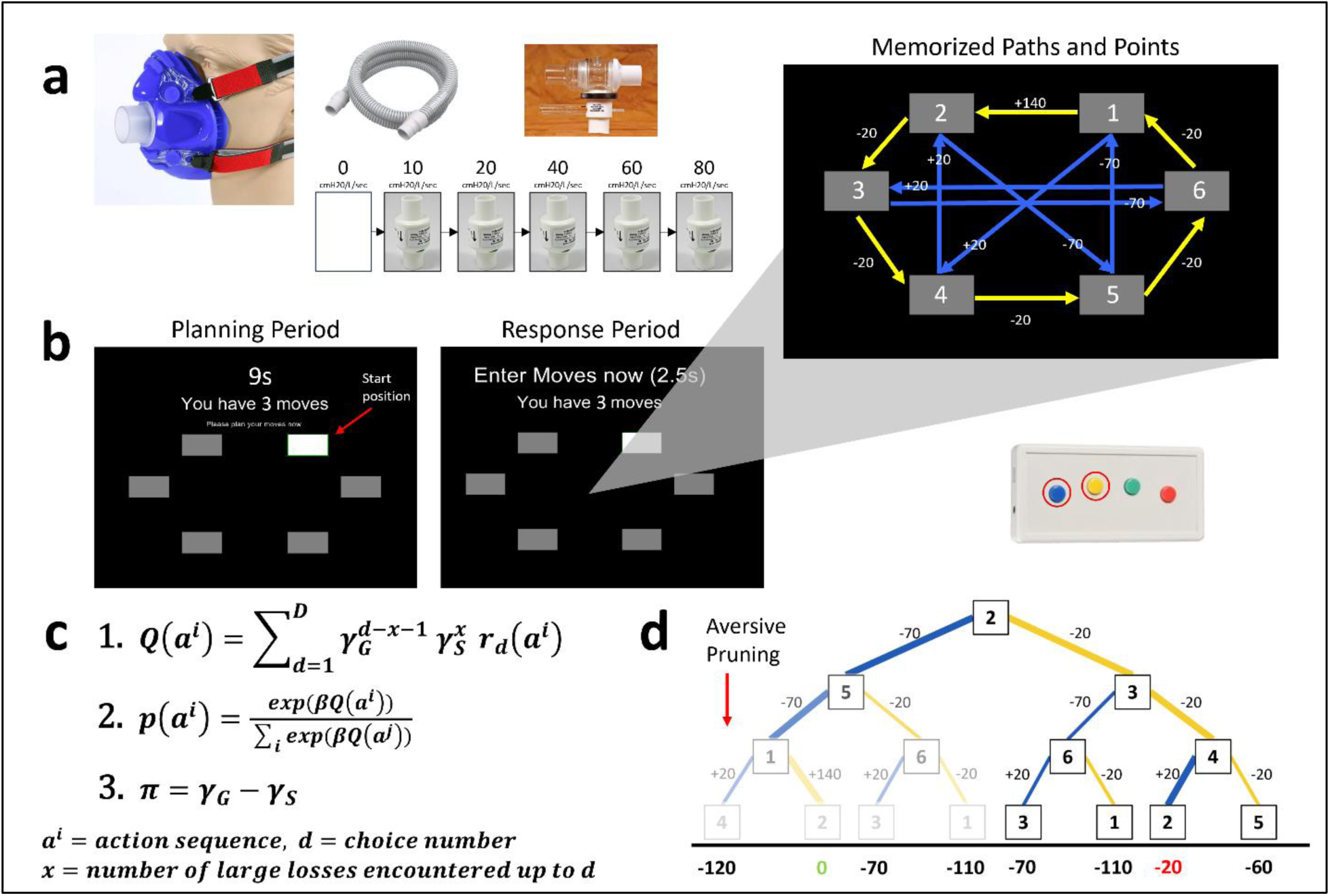
Study equipment, task interface, and computational model. **A)** Equipment used for anxiety induction: silicon mask with adjustable straps and single breathing port; resistors used to create resistance during inhalation and induce anxiety; two-way valve connected to the mask and ensures that inhalations engage one port while exhalations engage the other; tube connecting two-way valve to resistor. **B)** Graphical interface of the Planning Task. The blue button on the button box (center right) corresponds to transitions with blue arrows and the yellow button corresponds to transitions with yellow arrows. **C)** Computational model of 1) path valuation, 2) the probability of selecting a particular action sequence, and 3) calculation of **AP**. Note that, in the main text, **AP** = ***π***, **NLL-discounting** = **1** − ***γ_G_***, and **LL-discounting** = **1** − ***γ_S_***. Full model details are described in the Methods. **D)** Example decision tree based on an example starting position with point values for transitions and final path points demonstrating AP: namely, selectively discounting of further steps in a path that contains a large loss (i.e., options further down the tree depth are more faded/discounted than earlier steps). Points for the optimal path and the second-best path (indicated with thicker connecting lines) are shown in green and red, respectively. Colors of connections indicate whether the move was performed using the left (blue) button or the right (yellow) button.

Self-reported anxiety after each resistance level in the sensitivity protocol (i.e., testing how much initial exposure to each resistance level increased anxiety for each individual) is shown in the top panel of **Fig. 2**. Full results of models testing effects of group, resistance level, and their interaction on anxiety are reported in **Supplemental Materials**. Confirming prior results in an overlapping sample (31), anxiety levels increased with resistance, iMUDs had higher anxiety ratings than HCs, and anxiety increased more steeply in iMUDs than in HCs. Analogous figures and statistics for the other ratings given during the resistance sensitivity protocol (e.g., unpleasantness, fear, etc.) are provided in **Supplemental Fig. S1** and **Table S1**.

**Fig. 2.**
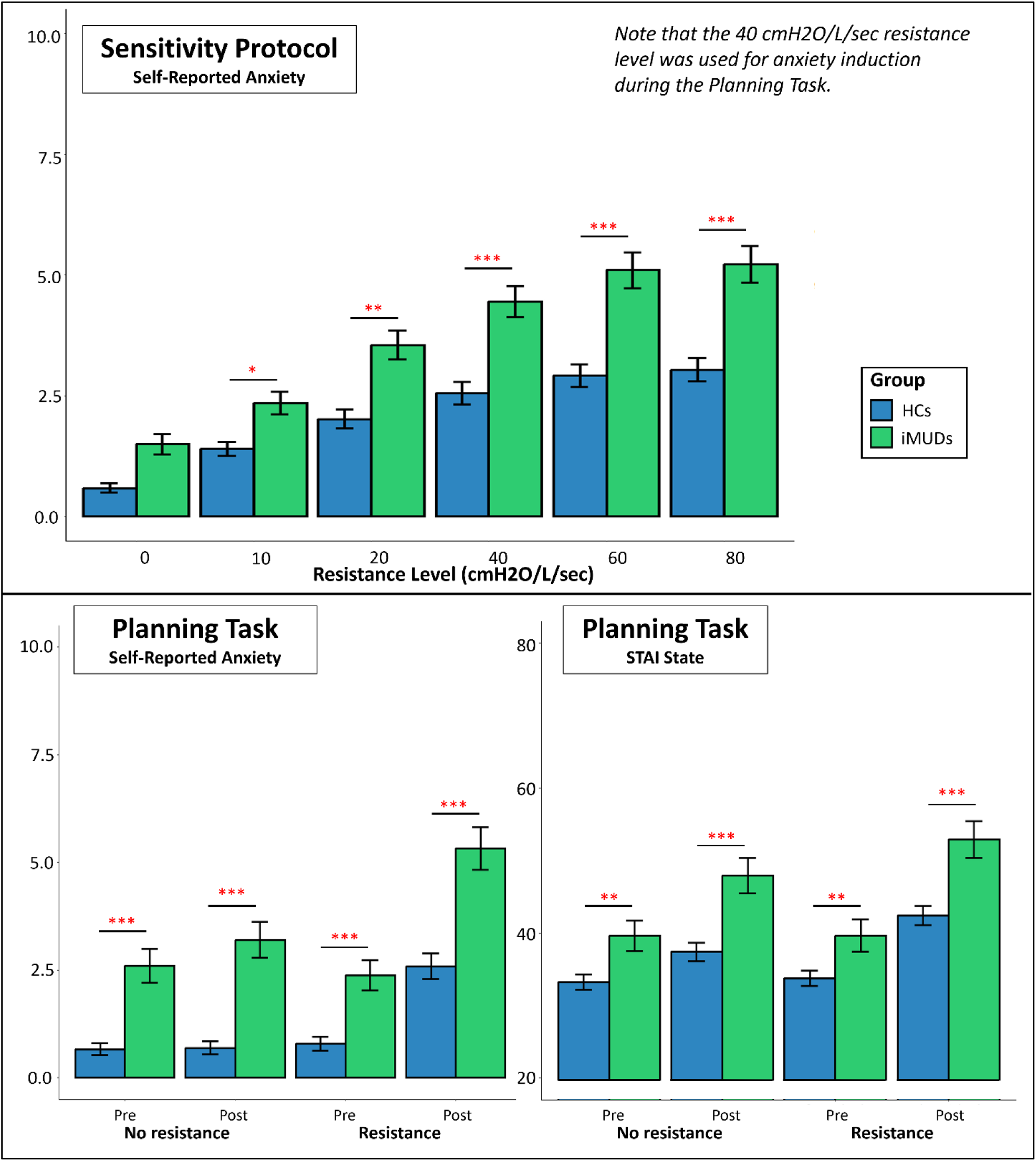
Anxiety induction efficacy. *Top*: Means and standard error bars for self-reported anxiety ratings across the resistance sensitivity protocol (scale 0-10). Anxiety for iMUDs (*n*=40) was higher than HCs (*n*=49; *F*(1,101)=14.21, *p*<.001, 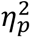=.12), and anxiety increased as a function of resistance level (*F*(1,977)=710.53, *p*<.001, 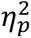=.42; b=0.644). *Bottom*: Means and standard error bars for self-reported anxiety (scale 0-10) and State-Trait Anxiety Inventory (STAI; (32)) State ratings from pre- to post-task for runs with and without the breathing resistance. Again, anxiety was generally higher in iMUDs (*Fs*>14.39, *ps*<.001) and also increased with resistance level (*Fs*>4.50, *ps*<.001). Stars indicate significant differences in post-hoc comparisons between groups at each resistance level (top) or time point (bottom). *p<.05,**p<.01,***p<.001

During one run of task performance (counterbalanced order), participants were continuously exposed to a resistance level of 40 cmH2O/L/sec, chosen to maintain a moderate (but tolerable) anxiety level (i.e., based on previous work using this paradigm (33) and confirmed by results of the sensitivity protocol). The other run was completed with no resistance. Anxiety ratings (both self-reported and from the State-Trait Anxiety Inventory [STAI] State scale) gathered before and after each run of the task are shown in the bottom panel of **Fig. 2**. Full results for analogous models testing effects of group, time (i.e., *pre* or *post*), and resistance condition (and possible interactions) are reported in **Supplemental Materials**. Overall, these results indicated that the aversive state induction protocol was effective at increasing anxiety levels.

### Aversive pruning was elevated in individuals with methamphetamine use disorder

Computational models with and without aversive pruning were compared using the Bayesian Information Criterion (BIC; shown in **Supplemental Fig. S2**). This confirmed that the aversive pruning model best explained participant behavior (ΔBICs≥2323 indicating very strong evidence). All analyses that follow were restricted to the fit parameters included in this model. Values in the present sample for each computational parameter demonstrated sufficient normality under both resistance conditions (skew<|2|; density plots shown in **Supplemental Fig. S3**).

The behavioral task completed by participants in this study was a version of the Sequential Planning Task described in Huys, Eshel (30). Participants first underwent extensive training to learn available action options in the task (i.e., transitions between different nodes [gray boxes]) and associated point values (i.e., values of −20, +20, −70, or +140 associated with each possible transition). In the task, they were asked to plan sequences of 3, 4, or 5 moves through a graph with 6 nodes on each trial (see **Fig. 1B)**. The possible transition options and point values were not shown during the task and needed to be drawn from memory. The starting node differed on each trial. Moves were planned during a 9-second “planning period” and then entered during a 2.5-second “decision period,” with the goal of maximizing points won on each trial.

To verify adequate recall of transitions and associated point values, a subset of participants (HCs=46, iMUDs=36) also completed a post-task assessment (**Supplemental Fig. S4**). Overall accuracy on the post-test confirmed successful retention of transitions and point values (M=87.4%, SD=19.3%). However, there was a significant difference between the two groups on overall accuracy (*t*(80)=4.95, *p*<.001, Cohen’s *d*=1.10), as well as for a derived accuracy metric linked to aversive pruning, reflecting the ratio of accuracy on questions involving transitions with large losses (−70 pts) compared to all other questions (*t*(80)=2.26, *p*=.027, Cohen’s *d*=0.50). In both cases, HCs had higher accuracy than iMUDs (HC: average overall accuracy=96%, average derived metric accuracy=98%; iMUDs: average overall accuracy=77%, average derived metric accuracy=80%). Thus, in relevant analyses below, we confirmed whether differences in any computational measures could be accounted for by these memory differences (while noting that aversive pruning-like behavior reduces the number of times that large-loss transitions were observed during the task, which would itself be expected to lead to worse post-task memory).

Model parameter values by group and resistance condition (as well as model-free measures of behavior) are presented in **Table 1** (left) and visualized in **Fig. 3**. Results of the *group* effect in each LME are also shown in **Table 1** (right); all other statistical results are provided in **Supplemental Table S2A**. The *Resistance* effect was not significant in any model (*Fs≤*3.68*, ps≥*.058).

**Fig. 3.**
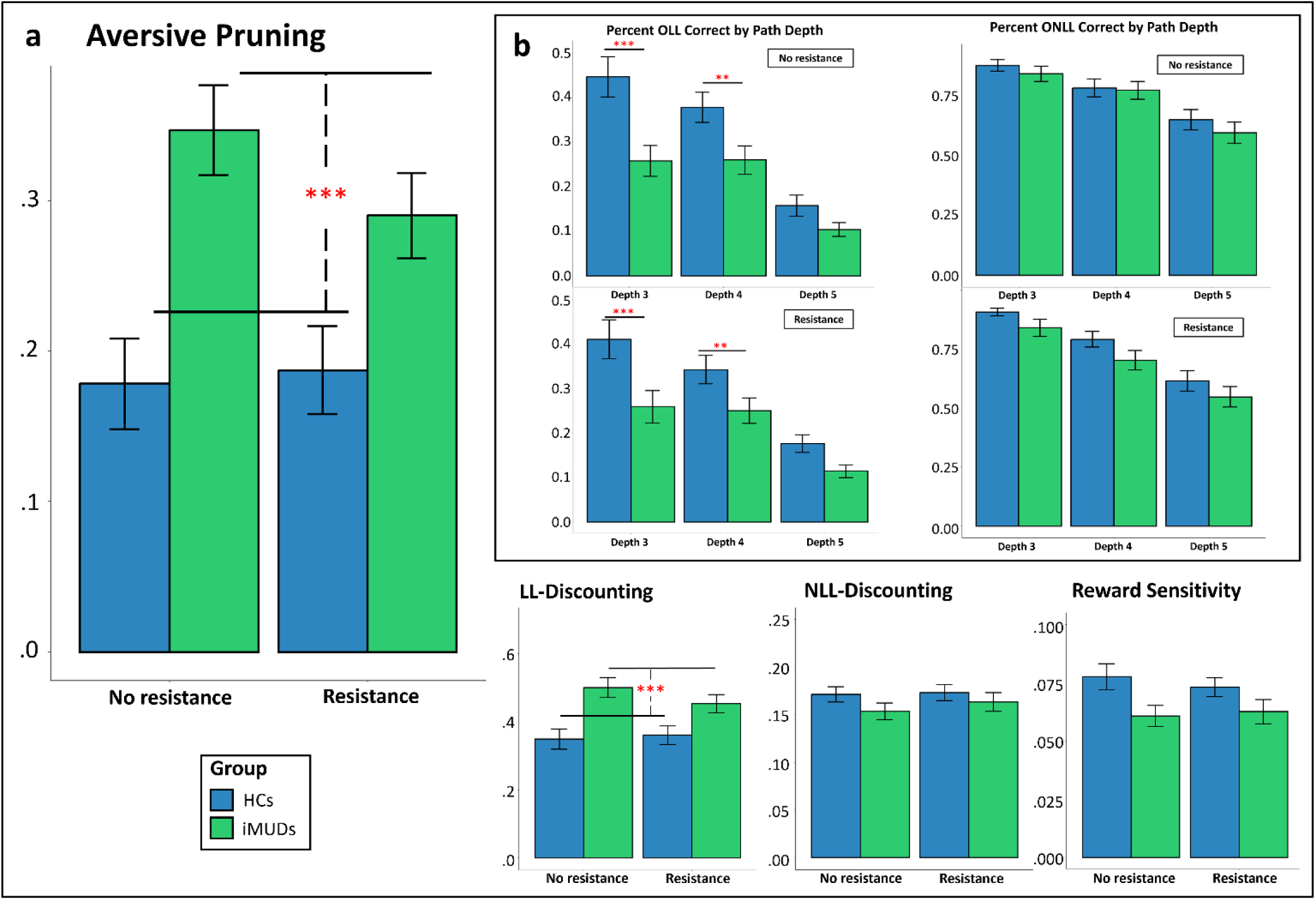
Computational parameters and model-free metrics of behavior. **A)** Model parameter means and standard errors separated by group and resistance condition (iMUDs: *n*=40; HCs: *n*=49). Independent of resistance level, iMUDs had larger **AP** estimates (*F*(1,100)=16.46, *p*<.001, 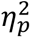=.14) and larger **LL-discounting** estimates (*F*(1,100)=13.45, *p*<.001, 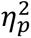=.12) than HCs. **B)** Choice accuracy differed by group in trials where the optimal path included large losses (OLL trials; *F*(1,490)=25.30, *p*<.001, 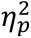=.05), driven by differences at depths 3 and 4. Stars indicate significant effects. LL=large loss; NLL=no large loss. \**p*<.05, \*\**p*<.01, \*\*\**p*<.001

**Table 1.**
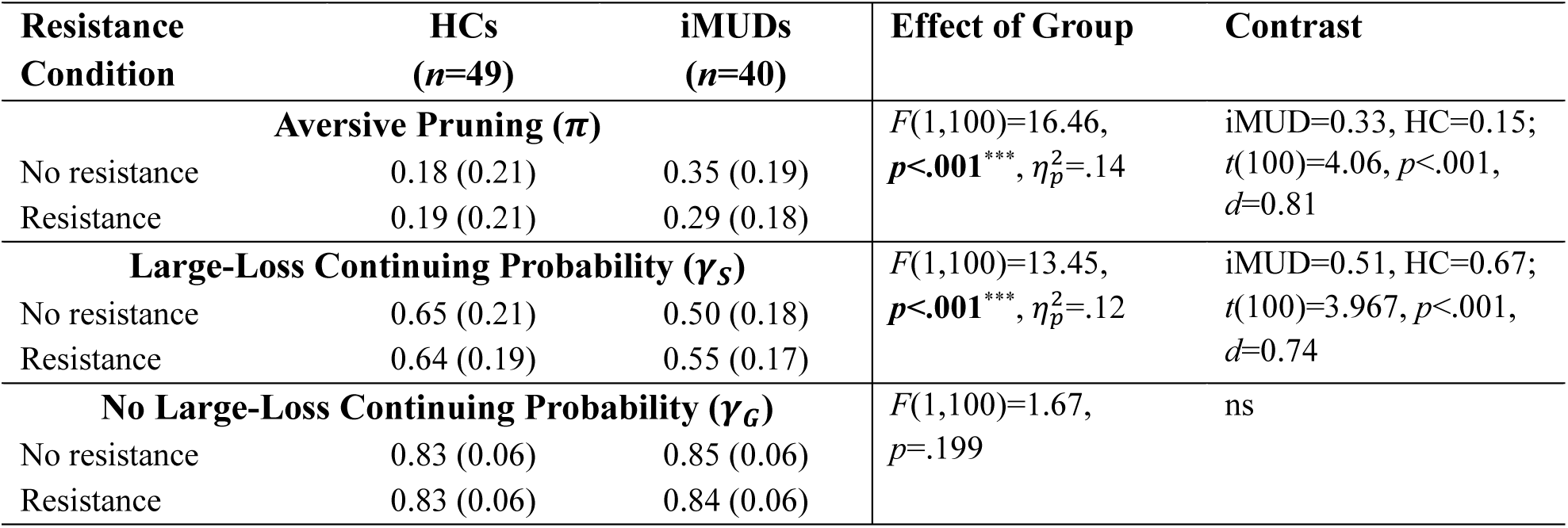
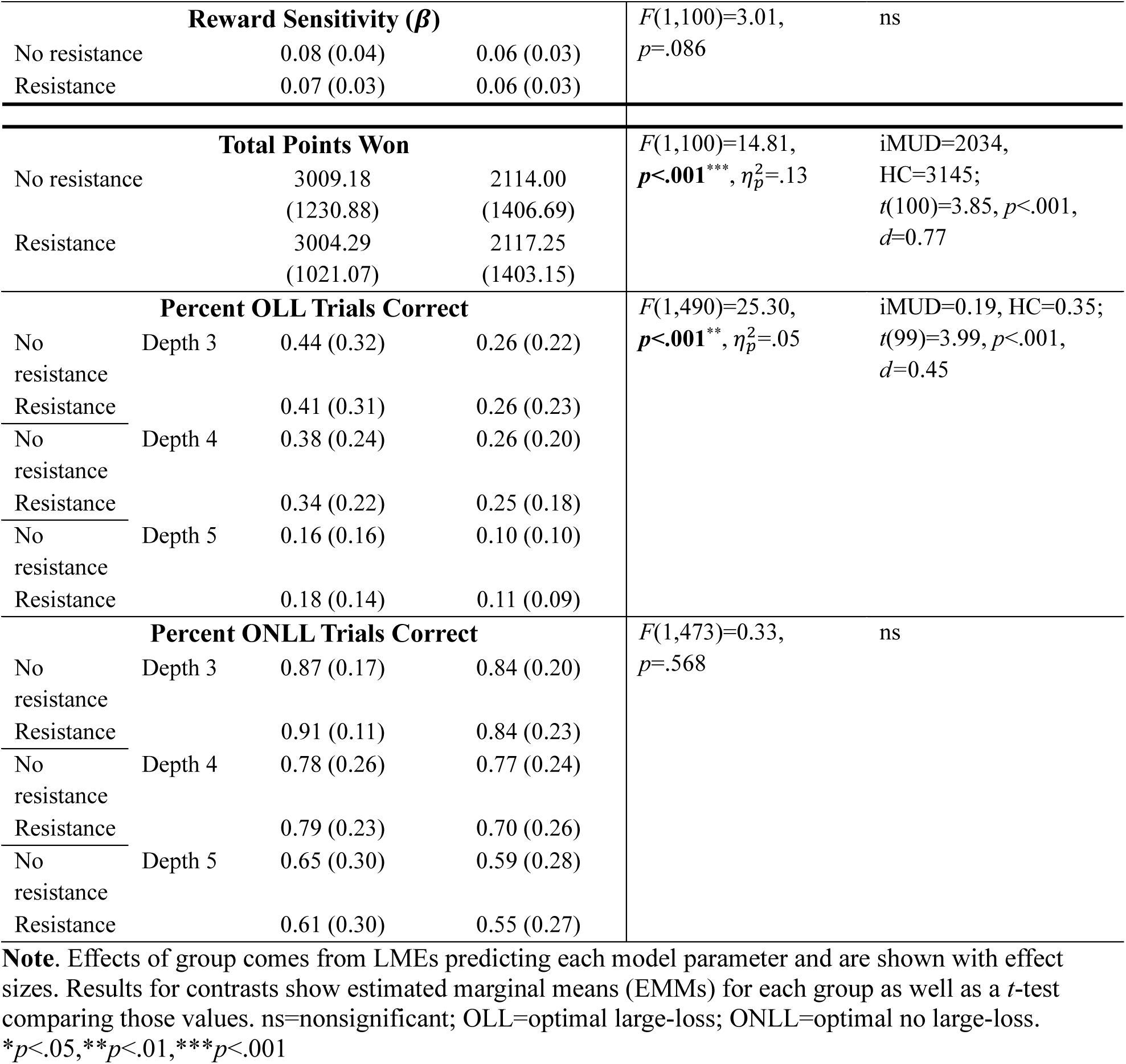
(Left) Computational model parameters and model-free task metrics by group and resistance condition. (Right) Statistics for the *group* effect in LMEs predicting each behavioral metric based on group, resistance condition, and their interaction. Post-hoc contrasts are also shown for significant effects.

With respect to model parameters, we first tested for effects on aversive pruning (**AP**), which reflects the difference between the probability that individuals will consider plans with large losses (**LL-discounting)** compared to when there are no large losses (**NLL-discounting**). In a model predicting **AP**, with group, state anxiety ratings, and their interactions with resistance condition as predictors, while accounting for age and sex, there was a significant effect of group (*F*(1,100)=16.46, *p*<.001, 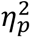=.14), such that iMUDs pruned more than HCs (large effect size of Cohen’s *d*=0.81 in post-hoc contrasts). There was also a significant *Group x Resistance* interaction (*F*(1,90)=5.17, *p*=.025, 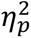=.05) indicating that iMUDs pruned more without the added resistance than with it. Finally, an effect of sex was also observed (*F*(1,84)=5.98, *p*=.017, 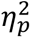=.07), indicating greater pruning in female participants.

For further interpretation, we subsequently analyzed **LL-discounting** and **NLL-discounting** separately. When predicting **LL-discounting** in analogous LMEs, there were again main effects of group and sex, such that iMUDs discounted more than HCs and that female participants discounted more than male participants (*F*(1,85)=4.70, *p*=.033, 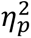=.05). Similarly, there was a significant *Group x Resistance* interaction (*F*(1,90)=4.84, *p*=.030, 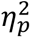=.05), reflecting a pattern consistent with what was found in models of **AP**.

Analogous LMEs predicting **NLL-discounting** showed no significant effects (*Fs*≤2.38, *ps*≥.126), suggesting findings for **AP** were explained by differences in **LL-discounting**. In other words, the alternative (or complementary) hypothesis that iMUDs would show a shorter planning horizon in general was not supported.

The other possible mechanistic explanation of planning deficits in iMUDs was a reduction in reward sensitivity (**RS** [*β* in model equations]; i.e., the degree to which expected overall reward differences between paths guided choice). Here, LMEs analogous to those above did not show any significant results, suggesting **AP** differences offered the primary explanation. Upon visual inspection, this was somewhat surprising given the notably greater **RS** values (numerically) in HCs (see **Fig. 3**). Further investigation revealed that when we removed state anxiety as a predictor (i.e., which also differed between groups), a significant group effect was revealed consistent with the apparent difference (*F*(1,85)=5.65, *p*=.020, 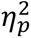=.06); thus, covariance between anxiety levels and group may have masked this effect.

### Group differences in task performance were primarily at shorter depths

After assessing model-based behavior, we also performed complementary assessment of model-free metrics (i.e., overall points won and accuracy by path depth on trials with and without large losses on the optimal path [OLL and ONLL, respectively]). Bar plots for accuracy by trial depth and resistance condition are shown in **Fig. 3**. Here we observed a main effect of group on overall points won, such that HCs won more points than iMUDs (see **Table 1**). There was also a significant effect of sex (*F*(1,85)=5.04, *p*=.027, 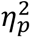=.06), such that male participants scored higher than female participants (*t*(85)=2.25, *p*=.027, Cohen’s *d*=0.49).

When predicting percentage of correct OLL trials by path depth, there were effects of group (see **Table 1**), depth (*F*(1,437)=158.79, *p*<.001, 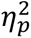=.27), and their interaction (*F*(1,437)=11.84, *p*<.001, 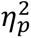=.03). Post-hoc contrasts showed that: 1) HCs had higher overall accuracy than iMUDs; 2) accuracy decreased as path depth increased (b=-0.102); and 3) OLL accuracy decreased more sharply in HCs (ET=-0.13) than iMUDs (ET=-0.07; *t*(437)=3.44, *p*<.001, Cohen’s *d*=0.33). This last effect was due to the fact that HCs were more accurate than iMUDs in depth 3 and depth 4 but dropped more steeply to become equivalent to iMUDs in depth 5. There was also an effect of sex (*F*(1,85)=10.06, *p*=.002, 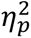=.11), where male participants (EMM=0.33) had higher OLL accuracy than female participants (EMM=0.21; *t*(85)=3.17, *p*=.002, Cohen’s *d*=0.70).

For percentage of correct ONLL trials, there was a main effect of depth (with greater path depth predicting worse accuracy; *F*(1,437)=215.49, *p*<.001, 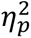=.33; b=-0.132). No other significant effects were observed (*Fs*≤2.44, *ps*≥.119).

### Group differences were not explained by memory, anxiety/depression, or comorbidities

Results of all LMEs above were largely equivalent when including working memory and post-task memory scores as additional covariates (i.e., in the subset of participants with available data; detailed results provided in **Supplemental Table S2B**). In particular, all observed group differences (apart from those noted for **RS**) remained significant.

No models testing possible effects of trait anxiety and depression (i.e., scores on the Overall Anxiety Severity and Impairment Scale [OASIS; (34)] and the Patient Health Questionnaire [PHQ; (35)], respectively) showed significant results (see **Supplemental Tables S3A** and **S3B**). When comparing iMUDs with and without specific comorbid substance use diagnoses, we found no differences in **AP** estimates between those with and without alcohol or opioid use disorders (noting that we were 80% powered to detect large effects only; 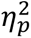>.17). However, we did observe that individuals with alcohol use disorder showed lower **LL-discounting** than those without (see **Supplemental Materials** for details; see **Supplemental Table S4** for a full breakdown of comorbidities). However, values for both alcohol (37.5%) and non-alcohol users (62.5%) each remained numerically higher than HCs (alcohol: M=0.41±0.16; non-alcohol: M=0.52±0.14; HCs: M=0.36±0.18). No other significant differences were found for any computational parameter.

Due to the difference in sex ratios between groups, supplementary models also tested for potential *Group x Sex* interactions to evaluate whether group differences in **AP** were confounded by sex. Results did not show a significant interaction and confirmed that the group difference was present in both sexes. Secondary analyses showed similar non-significant interactions when predicting, **RS**, total points won, or percentage of correct ONLL trials. However, we did note significant *Group x Sex* interactions suggesting that: 1) group differences were larger in males for **LL-discounting** and percentage of correct OLL trials; and 2) **NLL-discounting** estimates were selectively lower in iMUDs than HCs in females (see **Supplemental Fig. S5**).

We also examined length of abstinence (measured in days since last methamphetamine use: M=57.25±42.65), days since start of treatment (M=30.28±11.00), and medication status (*n*=30 medicated) as possible predictors of model parameters in iMUDs alone. Results of these tests are detailed in **Supplemental Materials**. No significant effects were found (*Fs*≤3.33, *ps*≥.077).

Although resistance order was counterbalanced across all participants, we also checked for potential effects of condition order. Briefly, results for **AP** and **LL-discounting** were unchanged. In contrast, **NLL-discounting** estimates for HCs (but not iMUDs) decreased significantly from run-1 to run-2, **RS** increased from run-1 to run-2 across participants, and HCs showed higher **RS** values than iMUDs only in those who underwent the breathing resistance during run-1 (see **Supplemental Fig. S6** and **Supplemental Table S5**).

While **AP** estimates were higher in iMUDs, it was unclear whether this should be viewed as maladaptive. To evaluate this interpretation, model parameters were correlated with model-free metrics of task performance. Here, all relationships were in expected directions (**Fig. 4A**). Most importantly, **AP** showed a linear, negative relationship with total points won (*rs*≥|.26|, *ps*≤.013), indicating that task performance was worse in participants who engaged in more pruning. Additionally, **RS** positively correlated with total number of points won, and **AP** negatively correlated with percentage of correct responses on trials for which the optimal path included a large loss (OLL trials).

**Fig. 4.**
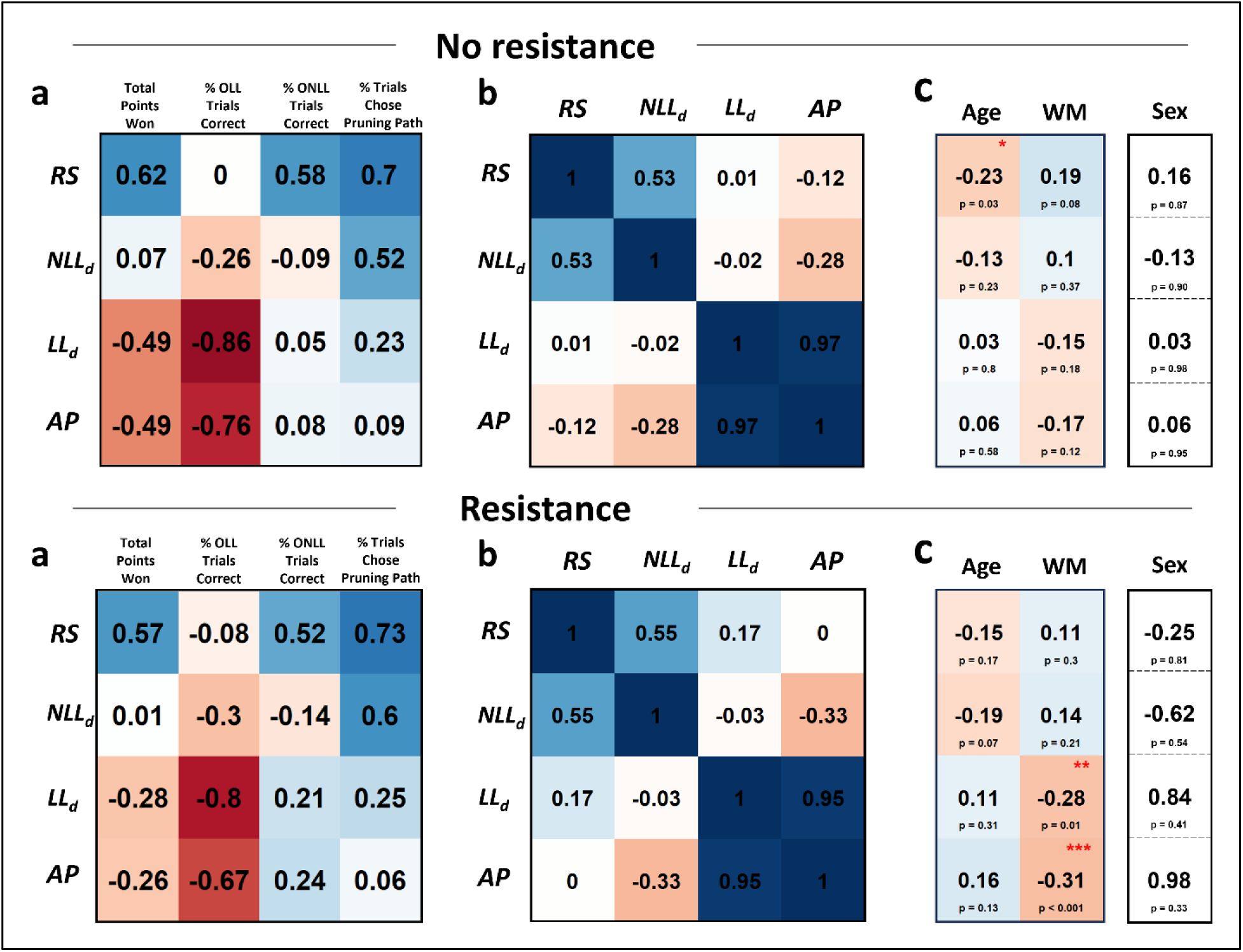
Computational parameter correlates. **A)** Correlations between model parameters and model-free metrics across groups. **B)** Inter-correlations between model parameters. **C)** Correlations between model parameters and covariates (statistics shown for sex represent t-values from independent samples t-tests, where negative signs indicate greater values in male participants). All relationships are shown separately for parameters under no resistance (top) and resistance (bottom) conditions. *OLL*=trials in which optimal path contains a large loss, *ONLL*=trials in which optimal path contains no large loss. *RS*=Reward Sensitivity, *NLL_d_*=ONLL Path Continuing Probability, *LL_d_*=OLL Path Continuing Probability, *AP*=Aversive Pruning.

Inter-correlations between the three model parameters for all participants demonstrated sufficient differentiability (**Fig. 4B**). Exploratory relationships between model parameters and demographic variables are shown in **Fig. 4C**.

### Craving and withdrawal symptoms relate to both resistance sensitivity and model parameters

When restricting to iMUDs (*n*=40), linear models (LMs) predicting resistance sensitivity (i.e., change in anxiety level from pre- to post-sensitivity protocol session), using DAST (drug abuse), MAWQ (withdrawal), and DSQ (craving) scores showed largely nonsignificant results (*Fs*≤1.90, *ps*≥.177; details in **Supplemental Materials**). However, more severe MAWQ emotional symptoms were associated with greater increases in anxiety after the sensitivity protocol (*F*(1,35)=9.15, *p*=.005, 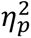=.21, b=0.602; see **Fig. 5a**), which remained significant after correcting for four subscale comparisons. No other significant predictors of changes in anxiety ratings (measured by self-reported anxiety and STAI State) from pre- to post-task were found (*Fs*≤3.96, *ps*≥.054).

**Fig. 5.**
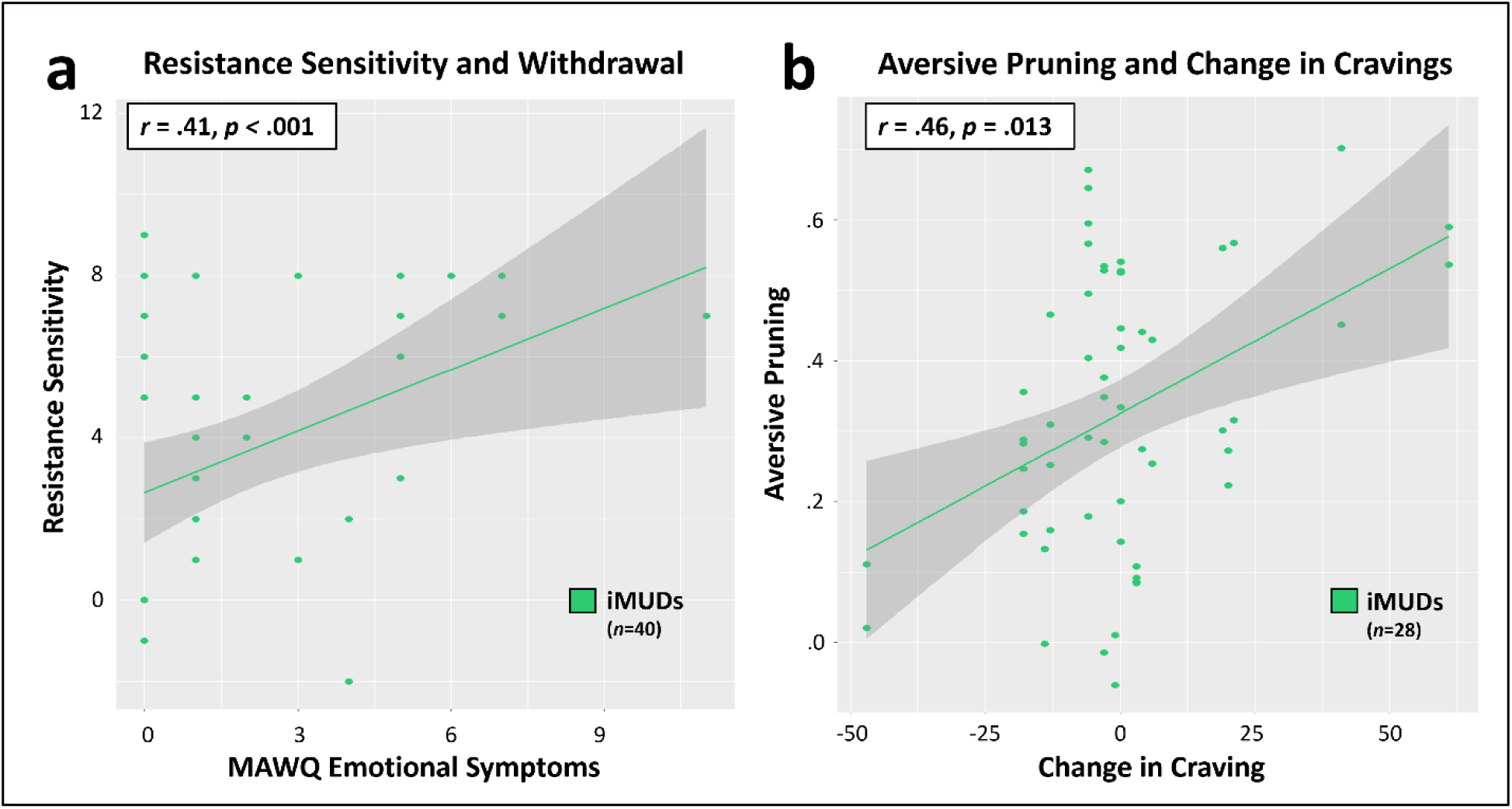
Symptoms relating to anxiety and parameters. **A)** Association between withdrawal symptoms and resistance sensitivity (as measured by change in self-reported anxiety ratings from pre- to post-resistance sensitivity protocol) in iMUDs (*n*=40; *r*=.41, *p*<.001). **B)** Association between change in craving (as measured by the DSQ from before to after completion of the anxiety induction) and aversive pruning (**AP**; averaged across resistance condition) during the Planning Task in iMUDs (*n*=28; *r*=.46, *p*=.013). This indicated that those with greater aversive pruning scores also had the greatest increase in craving in response to the anxiety induction (averaged across resistance conditions).

In an LME predicting **AP** in iMUDs, including DAST scores, resistance, and their interaction, and accounting for age and sex, there was a significant main effect of DAST scores (*F*(1,36)=12.15, *p*=.001, 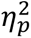=.25), indicating (surprisingly) that more severe consequences of drug use were associated with less pruning (b=-0.043). Analogous models replacing DAST scores with each MAWQ scale (separately) found no effects of withdrawal symptoms on **AP** (*Fs*≤1.28, *ps*≥.266). In a model including baseline craving symptoms (DSQ; *n*=28), there was no significant effect of craving (*F*(1,24)=2.74, *p*=.111). However, changes in DSQ scores after anxiety induction, accounting for baseline craving scores and the interaction between DSQ change and resistance condition, showed a positive association with **AP** (*F*(1,23)=8.82, *p*=.007, 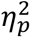=.28), indicating that an increase in craving following anxiety induction (i.e., within the pre-task resistance sensitivity protocol) was associated with more pruning (b=0.005; **Fig. 5b**). There was also a significant negative effect of baseline craving on **AP** (*F*(1,23)=5.70, *p*=.026, 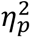=.20), indicating that higher baseline craving predicted less pruning (b=-0.002).

To understand the unexpected negative relationships between **AP** and baseline DSQ and DAST scores, we looked at individual scale items as predictors in LMEs that included resistance condition only. We found that a small number of items reflecting self-control on both measures acounted for these relationships (DAST items 3, 4, 8; *Fs*≥5.40, *p*s≤.026 [N=40]; DSQ items: 2, 14, 15, 27, 30, 36; *Fs*≥4.34, ps≤.047 [N=28]), highlighting a context in which pruning can be adaptive within iMUDs (e.g., individuals with greater pruning were more likely to say no to items such as “Have you engaged in illegal activities in order to obtain drugs?” that involve possible negative immediate outcomes).

Results of analogous models testing effects of symptoms on other model parameters are in **Supplemental Materials**. Briefly, results found for **LL-discounting** matched those of **AP**. When predicting **NLL-discounting**, there was also an interaction between functional (MAWQ) withdrawal symptoms and resistance condition (*p*=.047), suggesting that anxiety induction increased the effect of withdrawal state on planning horizon.

### Cognitive reflectiveness partially explained group differences in aversive pruning

As expected, an LME including Cognitive Reflection Test (CRT) scores (*n*=88), resistance condition, age, and sex as predictors revealed higher cognitive reflectiveness tendencies were associated with less pruning (*F*(1,84)=17.43, *p*<.001, 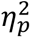=.17, b=-0.036). CRT scores similarly showed a negative relationship with **LL-discounting** (*F*(1,84)=17.60, *p*<.001, 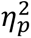=.17) and a positive relationship with **RS** (*F*(1,84)=12.38, *p*<.001, 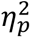=.13), but were not predictive of **NLL-discounting** (*p*=.698). We also confirmed these relationships in iMUDs alone and observed the same pattern of results for **AP** (*p*=.009, 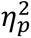=.18), **LL-discounting** (*p*=.013, 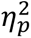=.16), and **NLL-discounting** (*p*=.422), while the effect on **RS** was no longer significant (*p*=.334). Relationships remained the same if accounting for working memory and post-test accuracy (in the smaller sample with available data), with the exception of the effect on **AP** in iMUDs alone, which, despite similar effect size (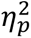=.13), rose slightly above the threshold for significance (*p*=.054; see **Supplemental Materials**).

These results suggested a potential mediation model in which greater **AP** in iMUDs might be explained by lower CRT scores. As shown in **Fig. 6**, testing this model revealed a significant indirect effect (*ab*=.063, *p*=.026, 95% CI: [.01,.13]), as well as a significant direct effect (*c*=.111, *p*=.024, 95% CI: [.01,.21]), indicating partial mediation (total effect *c*′=.174, *p*<.001, 95% CI: [.09,.25]). Thus, group differences in **AP** were partially accounted for by variation in reflectiveness.

**Fig. 6.**
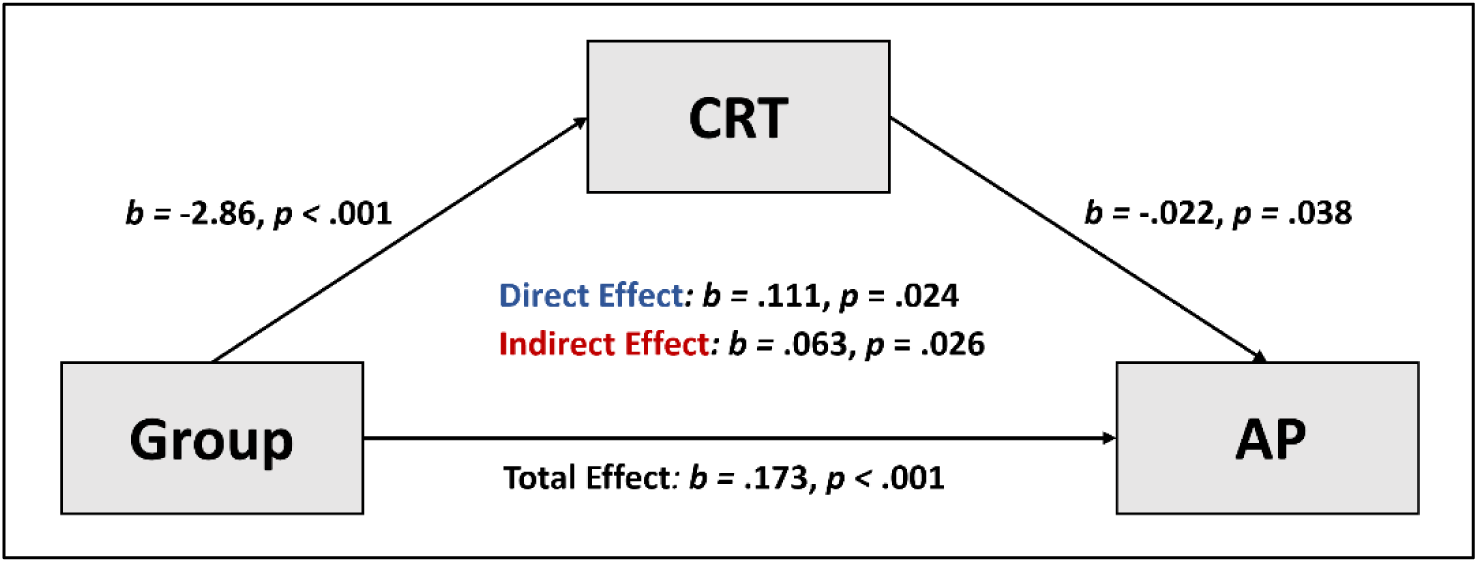
Mediation model. Graphical depiction of cognitive reflection as a significant partial mediator of group differences in aversive pruning (*n*=89). Note that the relationship shown between group and CRT accounts for age and sex.

We also assessed whether CRT scores related to craving in a way that could explain pruning differences in iMUDs. In those with available data, we found that both changes in craving scores after anxiety induction and DAST scores each showed noteworthy, but non-significant, trends with CRT (DSQ: *F*(1,24)=4.06, *p*=.055, 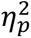=.14; DAST: *F*(1,37)=3.54, *p*=.068, 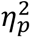=.09). There were no relationships observed with withdrawal symptoms (*Fs*≤0.43, *ps*≥.517).

Finally, as a supplementary exploration of potential relationships between model parameters and impulsivity/reward-seeking, we performed a latent factor analysis across several available measures from the larger study and tested relationships with resulting factor scores. The details of this analysis are described in **Supplemental Materials**. In brief, we did not find evidence for any relationships between factor scores for impulsivity or reward-seeking and model parameters within iMUDs. Descriptive information and group comparisons for included measures are shown in **Supplemental Table S6**.

## DISCUSSION

This study evaluated computational mechanisms of multi-step planning in individuals with methamphetamine use disorder (iMUDs) and healthy comparisons (HCs), and tested effects of an aversive interoceptive state. Computational measures included aversive pruning (**AP**; avoiding plans with large short-term losses), planning horizon (number of future steps one considers), and reward sensitivity (the degree to which planning is guided by expected reward). We observed substantially greater **AP** in iMUDs compared to HCs, independent of affective state, but no difference in overall planning horizon. To our knowledge, no previous study has examined this effect. Interestingly, group differences in **AP** were also partly mediated by cognitive reflectiveness, and greater pruning further predicted greater increases in craving in response to aversive state induction. This may be especially important given that negative affective states are known to promote vulnerability to relapse (36, 37), which our results suggest could be amplified in individuals with greater pruning tendencies.

Contrary to expectations, higher **AP** within iMUDs was instead associated with *less* severe consequences of drug use. Item-wise analyses offered important insights here by highlighting ways in which more pruning may confer greater self-control in iMUDs and prevent them from taking actions with short-term negative outcomes (e.g., committing crimes to acquire drugs). Thus, while there is a stark elevation in **AP** in iMUDs overall compared to HCs, variation within iMUDs could have adaptive effects with real-world consequences. This emphasizes the context-specific nature of when AP should be expected to confer advantages vs. disadvantages.

Surprisingly, we found no evidence for greater pruning after aversive interoceptive state induction. In iMUDs, results actually suggested greater pruning at baseline. However, it should be noted that the induction protocol only generated modest changes in anxiety (i.e., ∼2 to 3 point increases on a 10-point scale). One possibility is that effects could be accounted for by known inverted-U relationships between arousal and cognition (38), in which the induction kept iMUDs in a more alert or concentrated state, and that greater anxiety would have been necessary to produce the opposite effect.

The relationship we observed between pruning and cognitive reflectiveness suggests that those who have developed the cognitive habit of “thinking things through” before making a decision may also be less susceptible to overuse of **AP**. This finding may relate to previous work demonstrating deficits in other prospective cognitive processes in methamphetamine users (e.g., prospective memory performance and directed exploration (39–41)). It is also builds on the larger body of work in computational psychiatry suggesting shifts from model-based to model-free control (for reviews, see (42, 43)).

Of potential clinical relevance, studies have demonstrated that reflectiveness can be improved with training (44–49). Thus, this could be a targetable mechanism through which **AP** might be reduced. In line with our present findings, it is also possible that those with stronger cravings during aversive states (e.g., stress, withdrawal) are those that become more short-sighted during decision-making, which could, in turn, promote relapse (36, 37). Future studies in larger samples should evaluate whether lower reflectiveness could link craving and pruning behavior, and whether interventions focused on increasing reflectiveness might reduce pruning and/or lessen chances of relapse.

Some important limitations and future directions should be considered. First, sex was imbalanced between groups. While we confirmed group differences were present for each sex separately, future work in a balanced sample should replicate these results. Some iMUDs also had comorbid disorders, but these comorbidities did not account for group differences. Available data to examine relationships between task behavior and craving were also limited; so these results should be seen as preliminary.

Task rewards were only associated with small monetary value, and did not have substance-relevant meaning. Assessment of pruning effects on tasks with substance-related rewards could be crucial to further understand how MUD affects planning, particularly in relation to craving (21). Future work might adapt this task to utilize drug-related cues as rewards and link task behavior to induced craving or other clinical outcomes.

As there were some differences in the surrounding study protocol for the two groups (see *Methods*), we also cannot rule out that this influenced behavior. It should also be highlighted that the design of the present study does not allow us to differentiate whether observed effects represent pre-existing vulnerability factors or effects of substance use itself. We did not find lower pruning in those with greater length of abstinence, but studies testing a wider range of abstinence periods will be important.

In summary, we found that individuals with methamphetamine use disorder exhibited elevated aversive pruning, compared with healthy participants, on a multi-step planning task designed to pit large anticipated losses in the short-term against optimal positive outcomes in the long-term. This novel finding suggests a model-based impairment in the ability to consider optimal plans that require one to endure short-term aversive states. This effect has potential real-world relevance, as it mirrors difficult decisions faced by this population in which pruning could maintain use (e.g., not being able to consider the long-term benefits of abstinance due to the anticipated short-term pain of withdrawal). It also highlights a potentially novel treatment target with correlates (i.e., reflectiveness) known to improve with training. If replicated in future work, crucial next steps will require longitudinal and intervention studies designed to assess how pruning might relate to vulnerability and treatment response, and whether it can be modified in a manner that could improve clinical outcomes.

## METHODS

### Participants

Data were collected at the Laureate Institute for Brain Research (LIBR) in Tulsa, Oklahoma. Eligible participants came from the Tulsa community, were 18-65 years old, weighed ≤250 pounds (due to equipment limitations), and did not have a history of traumatic brain injury or neurological disorders. Participants included those without any diagnosed psychiatric conditions or elevated symptom levels (HCs; *n*=49) and those diagnosed with amphetamine use disorder and methamphetamine as a primary drug of choice (iMUDs; *n*=40). Participants with MUD were recruited from recovery centers in the Tulsa area within 45 days of entry into treatment (demographic and symptom characteristics of this sample are shown in **Table 2**). A comorbidity breakdown for iMUDs is shown in **Supplemental Table S4**.

**Table 2.** Demographic makeup of both groups (mean and SD) and statistical tests to assess sample differences.

| Measure | Healthy comparisons<br>($n=49$) | Individuals with<br>methamphetamine use<br>disorder ($n=40$) | Statistical comparison |
| --- | --- | --- | --- |
| Sex (F) | 36 | 12 | $\chi^2(1) = 31.77, p < .001$ |
| **Age** | 35.14 (13.3) | 37.12 (7.12) | $t(87) = 0.85, p = .399$ |
| **Working Memory\*** | 52.19 (8.13) | 46.78 (10.03) | $t(82) = 2.73, p = .008$ |
| **PHQ-9** | 1.88 (2.39) | 4.28 (4.18) | $t(87) = 3.39, p = .001$ |
| **OASIS** | 1.18 (1.58) | 4.90 (3.53) | $t(87) = 6.61, p < .001$ |
| **DAST** | -- | 7.72 (1.91) | -- |

**Note.** Working memory scores come from the NIH Toolbox Cognition Battery. It is a fully corrected (age, sex, education, and race/ethnicity) measure from the List Sorting Test. PHQ-9=Patient Health Questionnaire (depressive symptoms; (69)), OASIS=Overall Anxiety Severity and Impairment Scale (34), DAST=Drug Abuse Screening Test (59). We used two-sided $t$ -tests (and a chi-squared test in the case of sex) to measure the significance of differences between the two groups.
\*Data only available in a subset of participants: HCs: $n=47$ , iMUDs: $n=37$ .

### Measures

#### Cognitive

##### List Sorting Working Memory Test

This assessment, from the NIH Toolbox Cognitive Battery, is a test of working memory (WM) that requires the responder to recall and correctly order visual and auditory stimuli (50). Scores on this assessment correspond to the total score of all items after being corrected for relevant demographic information (i.e., age, sex, education, and race/ethnicity; see Casaletto, Umlauf (51) for more details on these normative standards). A higher score on this assessment indicates greater WM capacity. As the planning task (**Fig. 1**) requires accurate memorization and recall of a complex graph, transitions, and associated point values, we included scores on this assessment to evaluate whether any differences found in task performance were not explained merely by WM.

##### Cognitive Reflection Test-7 (CRT)

In a 7-item extension of the original 3-item test (17), the CRT includes questions with intuitive, but incorrect, answers, where identifying the correct answers require a tendency to “stop and think” before trusting one’s initial responses. This therefore assesses an individual’s tendency to reflect before providing an answer (52). As poor planning, and aversive pruning in particular, could be attributed to less reflective tendencies, we included this measure to test this relationship.

#### Symptom Severity

##### Desires for Speed Questionnaire (DSQ)

This 40-item questionnaire, modified from the Desires for Alcohol Questionnaire, measures craving for amphetamine using a 7-point Likert scale indicating degree of agreement with each item (53). Because this measure was added part-way through data collection, only a subset of iMUDs (*n*=28) responded to the DSQ. This was assessed before and after anxiety induction in the resistance sensitivity protocol (see below) to measure the degree to which aversive state induction increased craving. We examined whether those with stronger changes in craving due to anxiety induction might also show greater planning dysfunction, as negative affective states very often precede relapse (54–56).

##### Methamphetamine Withdrawal Questionnaire (MAWQ)

The MAWQ (57) was adapted from the Amphetamine Withdrawal Questionnaire (58) to assess multi-dimensional symptoms of methamphetamine withdrawal. Based on 30 items, four sub-scores are extracted reflecting functional (e.g., appetite changes), physical (e.g., headaches), emotional (e.g., loss of interest or pleasure), and other symptoms (e.g., “I am not able to deal with stress as well as usual”). The MAWQ was included in this study as current withdrawal severity could relate to resistance sensitivity and potentially influence task behavior.

##### Drug Abuse Screen Test (DAST)

Created by Skinner (59), this test was designed as a brief (10-item) self-report instrument for indexing the degree to which substance use negatively impacts an individual’s life. Scores range from 0 to 10, with each item requiring a yes/no response and scores greater than 2 are considered evidence of substance abuse. Notably, this screening does not explicitly address alcohol or tobacco use.

##### State-Trait Anxiety Inventory (STAI)

This 20-item measure of anxiety levels contains language to target either state or trait anxiety (32). Scores range from 20 to 80 and state anxiety responses were collected before and after each run of the planning task in the present study.

##### Overall Anxiety Severity and Impairment Scale (OASIS)

This 5-item self-report measure assesses the degree to which anxiety interferes with standard daily functioning and quality of life (34). Scores range from 0 to 20, with scores greater than or equal to 8 successfully classifying (with ∼87% accuracy) individuals with an anxiety disorder (60).

##### Patient Health Questionnaire (PHQ)

Containing 9 items, the PHQ measures depressive symptoms in a self-report format (35). Scores of 5, 10, and 15 indicate mild, medium, and severe symptom levels.

Please note that, as part of a larger funded study, descriptive symptom severity data from these measures has previously been reported to characterize an overlapping sample (31). All analysis of this data in relation to computational task measures in the present report are novel.

### Experiment Design

#### Aversive state induction and resistance sensitivity protocol

Anxiety is often characterized by heightened emotional and physical arousal in response to a perceived threat, and previous literature points to a relationship between anxiety symptoms/disorders and perturbed breathing (for review, see Paulus (61)). In the present study, we attempted to induce a temporary state of interoceptive/somatic anxiety by altering breathing effort and producing feelings of air hunger. More particularly, participants were asked to breathe through a mask (**Fig. 1A**) that focused their inhalations and exhalations through a single breathing port. A two-way valve ensured that air was directed through different valves for inspiration and expiration. Resistors were added to the breathing port to adjust how difficult it felt to inspire by creating resistance during inhalation (in cmH2O/L/sec), while no resistance was applied to expiration.

As part of a larger study with other clinical groups, HCs completed the planning task inside an MRI scanner. To best match this environment, iMUDs completed the task inside a mock scanner designed to replicate the supine positioning and isolated/narrow space experienced while undergoing MRI. The study protocol was otherwise identical except that, due to scheduling constraints with the collaborating recovery homes, iMUDs completed all study activities in a single visit, while HCs completed some surveys and other study activities in one visit and performed the MRI session in a second visit on a different day). Before completing the task, all participants tested out the anxiety induction mask in the mock MRI scanner, designed to help people become accustomed to the breathing resistance in this environment. With the mask on (see **Fig. 1A**), participants were exposed to 6 increasing levels of resistance (i.e., 0, 10, 20, 40, 60, and 80 cmH2O/L/sec) for 60 seconds each and rated their anxiety immediately after each exposure:

> *“How much anxiety did you feel while breathing?”*(11-point scale; 0=no anxiety, 10=maximum possible anxiety one could tolerate)

Participants also provided ratings of other secondary questions pertaining to difficulty, valence, and arousal (see **Supplemental Materials**). This “resistance sensitivity protocol” allowed us to extract more granular estimates of sensitivity to the anxiety induction. Please note that anxiety ratings in response to this series of resistance levels have previously been described in conjunction with other data gathered as part of the larger study mentioned above (31). However, all analyses of this data in relation to computational measures described here are novel and focused on distinct research questions.

#### Planning task

The behavioral task completed by participants in this study was a modified version of the Sequential Planning Task described in Huys, Eshel (30). A visual representation of this task is shown in **Fig. 1B**. The player sees 6 squares with lines indicating unidirectional transitions from one square to another. Transitions also have associated point values, and the player learns these transitions and point values through extensive training and testing before playing the game (in which this information must be drawn from memory during planning). The objective in this task is to maximize points while transitioning between squares from the starting position (which varies trial-to-trial) using exactly the number of moves allowed in that trial. Moves are planned out during a 9-second “planning period” and then entered in sequence during a subsequent 2.5-second “response period”. In the present study, participants completed two runs of 72 trials (each run took just under 20 minutes) with allowed sequences varying from 3 moves to 5 moves on different trials (i.e., requiring the player to plan up to 5 sequential moves).

Given the apparent complexity and memory demand in this task, participants were required to complete a training on rules of the task in steps and includes a practice test. In order to pass the training, participants had to respond correctly to questions about transitions and point values to ensure adequate retention of the task structure and reduce effects of memory differences on performance. Additionally, participants completed a post-task assessment that evaluated the knowledge they retained of the possible transitions and associated points (see **Supplemental Fig. S3** for more details).

#### Procedure

As mentioned above, because data from iMUDs was collected as part of an internally funded study, while data collection from HCs was grant-funded as part of a larger study including MRI with other clinical groups, the protocol for the two samples differed slightly. For HCs, the planning task was completed on the second visit of a two-day study after they completed surveys and other activities on Day 1. On Day 2, after filling out initial screening questions and passing a urine drug analysis, HCs completed the task training and then performed the planning task inside an MRI scanner for their two task runs (fMRI data analysis for the larger grant-funded study is in progress and will be reported elsewhere). As described earlier, due to scheduling limitations with collaborating recovery homes, task training and performance for iMUDs were completed after a lunch break in a one-day study visit that began with the same surveys and other study activities that HCs performed on Day 1. Instead of the real MRI scanner, participants with MUD completed their two task runs in a mock MRI scanner to best match task environments for the two groups. Participants completed measures of self-reported anxiety and STAI State before and after each run of the task with a few minutes rest in between. Finally, after completing the planning task, retention of the transitions and associated point values was assessed in a post-test. However, this post-test was only added part-way into the study to help rule out potential retention-based confounds; thus, data for this measure was not collected in all participants (available data in HCs=46, iMUDs=36).

### Computational modeling and model fitting

Model-based behavioral analyses were performed using the computational modelling approach outlined in Lally, Huys (62) and conducted in MATLAB (R2022a). In brief, the value assigned to an action sequence, ***Q***(***a***^***i***^), was defined by the sum of rewards gained at each choice ***d*** in the sequence (denoted ***r_d_*(*a^i^*))**. In this model, a discounting parameter **0** ≤ ***γ*** ≤ **1** down-weights the influence of expected wins/losses at later steps in a given sequence. This parameter is separated into two independent components: 1) ***γ_S_*** refers to the degree to which paths that include large loss transitions (i.e., −70 points) are down-weighted as a function of depth (higher values indicate less discounting); and 2) ***γ_G_*** refers to the degree to which all other paths (i.e., including only combinations of −20, +20, and/or +140 points) are down-weighted as a function of depth (higher values indicate less discounting). Thus, the value function, as defined in this model, is as follows:

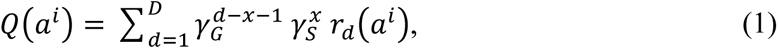

where *x* is equal to the number of expected large losses encountered up to point *d* in the path being considered. This has the effect of 1) effectively “turning on” ***γ_S_*** when ***x*** ≠ **0** (i.e., when a path has an expected large loss), and 2) otherwise exponentially decreasing the value of choices further down the decision-tree (by increasing the exponent on ***γ_G_***) to reflect nonlinear increases in discounting of later actions. Then, the probability of choosing a particular action sequence ***a***^***i***^ is:

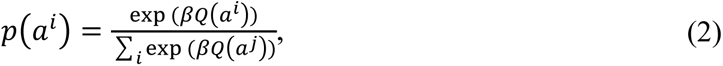

where an inverse temperature parameter ***β*** ≥ 0 (referred to here as *reward sensitivity*) scales the relative subjective value difference between sequences.

To assess an individual’s propensity to discount large loss branches above and beyond their general tendency to discount paths, we calculated a difference score, ***π*** = ***γ_G_*** − ***γ_S_***, corresponding to their *aversive pruning* (**AP**) value. Note that, for clarity, we report **1** − ***γ_G_*** (**NLL-discounting**) and **1** − ***γ_S_*** (**LL-discounting**) so that values correspond to the probability of discounting decision-tree paths.

The model fitting procedure used here is described in full detail in Huys, Eshel (30). In short, free model parameters (i.e., those that are fit to an individual’s choices; ***γ_S_***, ***γ_G_***, and ***β***) are found by maximizing the likelihood of observing the true set of all actions of a given participant (***A***_***i***_) under a set of parameter values (***h***) assuming Gaussian prior distributions ***p***(***h***|***θ*)** defined by parameters ***θ***. Together, this gives the following for the maximum posterior estimate ***m***_***i***_ for a participant:

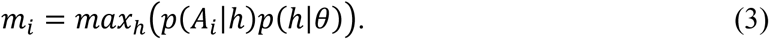

The prior distribution is set to be the maximum likelihood given all participant data in the present sample (including data under both task conditions). Maximization is achieved via Expectation-Maximization (63) with a Laplace approximation.

As in prior uses of this task, this aversive pruning model was compared to other simpler models that either: 1) removed ***γ_S_*** and assumed a single discounting term ***γ_G_*** was applied to all paths, or 2) included no discounting term and assumed the true summed value of all paths determined participants’ choices. The winning model was chosen via comparison of log-likelihoods (given all actions and parameters fit in each model) and the Bayesian Information Criterion (BIC), which provides a complexity cost favoring models with fewer parameters. For further details on these alternative models and comparison approach, see Huys, Eshel (30), Lally, Huys (62).

### Statistical analyses

#### Anxiety induction efficacy

All statistical analyses were performed using R Studio version 4.2.0 (64).

We first performed analyses to confirm the efficacy of the breathing resistance as an anxiety induction manipulation. Specifically, we ran LMEs (using Type II ANOVAs) with anxiety level as the outcome variable, and with group (sum-coded: HCs=-1, iMUDs=1), breathing resistance (sum-coded: no resistance=-1, resistance=1), and their interaction as predictors. This was done both for self-reported anxiety across resistance levels during the sensitivity protocol (coded as a continuous variable; resistance levels: 0, 10, 20, 40, 60, and 80 in cmH2O/L/sec) as well as self-reported anxiety and STAI state before and after the two task runs. To evaluate the possibility that the aversive state induced by the resistance had a relation to the negative state associated with craving, we also evaluated change in craving (i.e., DSQ score) before vs. after participants underwent the sensitivity protocol. Individual differences in this change in craving then provided a complementary metric of sensitivity to the manipulation.

#### Computational parameters and task behavior

We then performed a set of model validation analyses, including comparison to the other previously tested models mentioned above. Accuracy on the post-planning task memory assessment (i.e., *post-test*) was also checked both across and within groups. Specifically, we analyzed overall accuracy (mean and SD) and then ran a linear mixed effects model (using the *lmer* function within the *lme4* R package (65)) predicting response accuracy using question type (either testing memory of point values or transition directions), point value (−70, −20/+140, +20), and group as predictors (all categorical variables were sum-coded).

To address our primary aims of testing effects on model parameters, we then performed similar LMEs (using Type III ANOVAs) with group (sum-coded: HCs=-1, iMUDs=1), breathing resistance condition (sum-coded: no resistance=-1, resistance=1), and their interaction as predictors, while accounting for possible effects of age (centered) and sex (sum-coded: male=-1, female=1). These models also included an interaction between resistance condition and self-reported anxiety during the task to address the hypothesis that differences in heightened anxiety resulting from the breathing perturbation would impact task behavior.

Next, we performed comparable LMEs replacing model parameters with model-free metrics of behavior during the task (i.e., summary statistics describing task performance and choice behavior) to assess group differences and effects of resistance condition.

#### Secondary analyses of potential confounds

As a further check, analyses of task performance (both model-based and model-free metrics) were also repeated while accounting for working memory score and accuracy on the post-task memory test accuracy. These secondary analyses could only be performed in the subset of participants for which these data were available, but helped to confirm that differences were not explained by differences in either general cognitive ability or memory for different path outcomes. The post-task memory metric was calculated by taking the ratio of correct answers on the questions testing memory for large loss transitions (i.e., point value of, or transition through, a −70 path) to accuracy on all other questions. This measure was chosen to reflect aversive pruning behavior as closely as possible (i.e., in case pruning could be explained by worse memory for large-loss transitions). These secondary results are reported in **Supplemental Materials**.

Due to the sex imbalance in each of the two groups, we also tested for potential *Group x Sex* interactions in supplementary LMEs predicting each model parameter. This was done to confirm that any significant differences found were present in both males and females separately. We also examined possible effects of length of abstinence, days since start of treatment, and medication status in iMUDs for each model parameter. These LMEs (using Type III ANOVAs) also included state anxiety, resistance condition, its interaction with both previous variables, age, and sex.

Given that the clinical group had heterogeneous levels of depression and anxiety (comorbid diagnoses shown in **Supplemental Table S4**), we also subsequently verified observed relationships in models including anxiety (OASIS) and depressive (PHQ) symptoms as predictors (both centered). Significant effects were then interpreted via post-hoc contrasts using the *emmeans* package in R (66). In follow-up analyses, we also examined differences between those with and without specific co-morbid substance use diagnoses in the iMUDs where sample size allowed (i.e., alcohol use disorder and opioid use disorder; see **Supplemental Table S4**).

These models contained group designation (sum-coded: without disorder=-1, with disorder=1), breathing resistance condition, age, sex, and a *Group x Resistance* interaction.

We then examined correlations between model parameters and model-free behavioral measures to confirm expected relationships. Relationships between model parameters and demographic variables were also explored. All correlations were performed using the *corrplot* function (within the *corrplot* package (67)). Descriptive data for measures of interest are also displayed.

#### Clinical symptoms

To evaluate whether sensitivity to the anxiety induction might relate to disorder severity in iMUDs, we next ran linear models (LMs) with change in self-reported anxiety during the resistance sensitivity protocol (i.e., in response to the increase from 0 to 80 cmH2O/L/sec) as the outcome variable, and severity, withdrawal, or craving as predictor variables (i.e., DAST, MAWQ, or DSQ, each in separate models). To assess whether change in anxiety might also lead to changes in craving, the model with DSQ as a predictor also included change in DSQ from baseline to after the 80 cmH2O/L/sec resistance exposure. Baseline anxiety, age, and sex were also controlled for in each of these models.

Changes in anxiety during each task run were also assessed in a similar manner. Namely, using the same symptom measures above as predictors, we ran LMEs predicting individual differences in anxiety change (i.e., *post* minus *pre* for each task run), while accounting for effects of breathing condition, baseline anxiety levels, age, and sex.

#### Secondary dimensional analyses

We then evaluated whether individual differences in craving sensitivity – that is, changes in craving (DSQ) before vs. after the breathing manipulation – might relate to differences in aversive pruning. Supplemental analyses further explored the possibility of similar relationships with the other model parameters.

We also examined possible associations between **AP** and cognitive reflection. Specifically, we ran an LME including CRT scores as a predictor of **AP**. This model also included main effects of resistance interaction, age, and sex. An analogous LME was also run confirming results in iMUDs alone. Observation of a significant group difference in the initial LME led us to conduct a further mediation analysis testing CRT as a potential mediator of the relationship between group and **AP** (i.e., asking whether group differences in aversive pruning may be accounted for by group differences in cognitive reflectiveness tendencies). For this test, we used the *mediate* function (mediation package in R (68)) with 5000 simulations. This analysis was also repeated in **Supplemental Materials** to account for working memory.

Additional supplemental exploratory analyses were also performed in relation to available measures from the larger study reflecting impulsivity and reward-seeking. All details of these exploratory analyses are provided in **Supplemental Materials**.

## Data Availability

All data produced in the present study are available upon reasonable request to the authors.

## Acknowledgements

The authors would like to acknowledge Henry Yeh for assistance with statistical analyses.

## Conflict of interest or competing financial interests

The authors have no conflicts of interest to report.

## Author contributions

RS took the lead in designing and overseeing the study. CAL and RS played primary roles in planning and performing analyses and writing the manuscript. QJMH advised on analyses and edited the manuscript. SSK, JLS, and MPP aided in study design and execution and edited the manuscript. MMM, ST, and AEC assisted in analysis and data collection. All authors contributed to the writing and reviewing of the manuscript.

## Supplemental Materials

### Supplementary Methods

For exploratory tests of potential relationships with impulsivity and reward-seeking, the following additional measures were utilized in factor analyses (described below):

#### UPPS-P Impulsivity Scale (UPPS-P)

This self-report measure of impulsivity includes 59 items aimed to assess positive and negative urgency, (lack of) premeditation, (lack of) perseverance, and sensation-seeking (1). Of greatest interest was the Negative Urgency subscale reflecting an individual’s tendency to react impulsively as a result of intense negative affect. Scores on this scale were included in a factor analysis below, along with other dimensional measures, to evaluate their potential link to aversive pruning and other planning mechanisms.

#### Behavioral Inhibition System/Behavioral Activation System Scales (BIS/BAS)

These scales, comprising a measure typically examined together, contain items relating to inhibitory (BIS) and appetitive or approach (BAS) motivations (2). For our purposes, the BIS scale, which measures inhibition of behavior that may lead to aversive outcomes, was of primary interest. Here we specifically included BIS score in the impulsivity factor analysis as an additional potential predictor of aversive pruning and expected that greater inhibitory system activation would predict more aversive pruning.

#### Temporal Experiences of Pleasure Scale (TEPS)

This measure, created by Gard, Gard (3), captures anticipatory and consummatory experiences of pleasure in 18 self-report items. The TEPS subscales were included due to their potential connection to differences in reward sensitivity.

#### Additional ratings from resistance sensitivity protocol

In addition to self-reported anxiety levels, participants also gave responses to a series of other related questions immediately after exposure to each breathing resistance during the sensitivity protocol (exact wording listed below). Self-Assessment Manikin items assessed happiness and excitement (4) rated on a 5-point scale, while the others were rated similarly to self-reported anxiety (11-point scale). See **Figure S1** and **Table S1** below for associated visualization and statistics.

> “How calm or excited did you feel during loaded breathing?” (on the scale from 1 to 5)

> *“How happy or unhappy did you feel during loaded breathing?”* (on the scale from 1 to 5)

> *“How difficult did it feel to breathe?”* (0-no difficulty; 10-maximal difficulty you could tolerate)

> *“How much fear did you feel while breathing?”* (0-no fear at all; 10-maximal fear you could tolerate)

> *“How unpleasant did it feel to breathe?”* (0-not at all unpleasant; 10-maximal unpleasantness you could tolerate)

### Supplementary Analyses

#### Anxiety induction during sensitivity protocol and task

In a linear mixed effects model (LME) predicting anxiety levels during the sensitivity protocol, and including group, resistance level, and their interaction as predictors, all effects were significant (*Fs*≥14.21, *ps*<.001). Post-hoc contrasts revealed that: 1) the group effect was driven by higher anxiety in iMUDs (estimated marginal mean [EMM]=3.70) than HCs (EMM=2.09; *t*(87)=3.77, *p*<.001); 2) anxiety increased as resistance level increased (b=0.644); and 3) anxiety showed greater increases in iMUDs (estimated trend [ET]=0.79) than HCs (ET=0.49; *t*(443)=4.25, *p*<.001). Analogous results for other secondary self-report scales gathered during the sensitivity protocol session (e.g., subjective breathing difficulty, general unpleasantness, etc.) are available in **Supplemental Table S1**. These tended to follow a similar pattern as with self-reported anxiety.

For both measures of anxiety, group (*Fs*≥14.39, *ps*<.001), time (i.e., *pre* or *post*; *Fs*≥73.96, *ps*<.001), and resistance condition (i.e., somatic anxiety induction; *Fs*≥4.50, *ps*<.001) were significant predictors of anxiety. There were also significant *Time x Resistance* (*Fs*≥12.07, *ps*<.001) and *Time x Group* interactions (self-reported: *F*(1,259)=7.84, *p*=.005; STAI State: *F*(1,259)=10.35, *p*=.001), but no *Resistance x Group* or three-way interactions (*Fs*≤0.95, *ps*≥.332). Post-hoc contrasts revealed that: 1) iMUDs reported higher levels of anxiety than HCs (*ts*≥3.79, *ps*<.001); 2) anxiety at post*-*task was higher than anxiety at pre*-*task (*ts*≥8.80, *ps*<.001); and 3) anxiety ratings were higher for the *resistance* condition than the *no resistance* condition (*ts*≥3.79, *ps*<.001). Interactions indicated there was a significant increase in self-reported anxiety ratings from pre- to post*-*task performance in the *resistance* condition (*t*(259)=10.98, *p*<.001), but not in the *no resistance* condition (*t*(259)=1.48, *p*=.141). However, for STAI State, there were significant increases in anxiety ratings from pre*-* to post*-*task in both conditions (*ts*≥6.46, *ps*<.001). Finally, the *Time x Group* interaction reflected a larger difference in anxiety in iMUDs than HCs at post-task (although all post-hoc t-tests had *ps*≤.009).

#### Checking possible group x sex interactions

To ensure that the sex imbalance in the present sample was not explanatory of observed group differences, we conducted LMEs that included *Group x Sex* interactions predicting each model parameter and model-free metric. While the models predicting **AP**, **RS**, total points won, and percentage of correct ONLL trials revealed no significant interactions (*Fs*≤1.85, *ps*≥.177), the model predicting **LL-discounting** found that, in addition to the independent main effects of group and sex, there was a significant interaction (*F*(1,84)=4.47, *p=*.037, 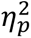=.05) indicating a difference between HC and iMUDs in male participants (*t*(84)=4.31, *p<*.001 where HCs had lower **LL-discounting** estimates) but not female participants (*t*(84)=1.39, *p=*.169). As additional checks, we also examined the other two model parameters and found another *Group x Sex* interaction when predicting **NLL-discounting** (*F*(1,84)=6.74, *p=*.011, 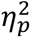=.07) such that a group difference was seen in only female participants (*t*(84)=2.91, *p=*.005 where female iMUDs had lower **NLL-discounting** estimates). Finally, the model predicting percentage of correct OLL trials saw results complementary to those for **LL-discounting** (*F*(1,84)=9.55, *p=*.003, 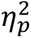=.10) where the group difference was present in male participants (*t*(84)=5.15, *p<*.001).

#### Effects of abstinence, treatment start, and medication

In models predicting each computational parameter in iMUDs alone (*n*=40), we included length of abstinence, days since treatment start, and medication status in separate LMEs along with their interaction with resistance condition, state anxiety and its interaction with resistance, age, and sex. When predicting **AP**, there was a marginal effect of medication status (*F*(1,35)=3.33, *p=*.077) where those taking psychotropic medication pruned more (EMM=0.35) than those who were not (EMM=0.24; *t*(35)=1.82, *p=*.077). Medication status was not predictive of any other parameter (*Fs*≤2.36, *ps*≥.133) and there were no effects of length of abstinence or days since treatment start for any computational parameter (*Fs*≤1.59, *ps*≥.215).

#### Diagnosis-specific analyses

In LMEs comparing iMUDs with vs. without opioid use disorder (OUD), there were no significant effects of or interactions with diagnosis for any model parameter (*Fs*≤2.53, *ps*≥.120). The same was true for most parameters when comparing those with vs. without alcohol use disorder (AUD), with the exception of **LL-discounting** (*F*(1,36)=4.82, *p=*.035, 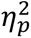=.12), which was significantly lower in those with AUD (EMM=0.40) than those without (EMM=0.51; *t*(36)=2.20, *p=*.035).

#### Dimensional relationships

##### Clinical symptoms

When restricting to iMUDs (*n*=40), a linear model (LM) predicting resistance sensitivity (i.e., change in anxiety level from pre- to post-sensitivity protocol session), using DAST scores and baseline anxiety levels as predictors, revealed a nonsignificant effect of DAST on sensitivity (*F*(1,35)=1.90, *p=*.177). Similar nonsignificant results were found when instead using MAWQ subscale scores as predictors (*Fs*≤0.80, *ps*≥.376) apart from emotional withdrawal symptoms (results shown in main text). To test if resistance sensitivity was predictive of changes in craving from pre- to post-sensitivity protocol, we included DSQ change scores (*n*=28), baseline anxiety, and baseline DSQ (along with age and sex) as predictors. The effect of DSQ change scores was not significant (*F*(1,22)=1.57, *p=*.223).

In an LME using DAST to predict change in anxiety pre- to post-task run (either self-reported anxiety or STAI State), accounting for pre-task anxiety before each task run, resistance condition, and the interaction between DAST and resistance condition, there were no significant main effects of symptom severity for STAI State or self-reported anxiety change (*Fs*≤3.52, *ps*≥.069). In analogous models replacing DAST with each MAWQ scale, no scale showed significant effects for either anxiety metric (*Fs*≤3.96, *ps*≥.054). Finally, when looking at only iMUDs with available DSQ data (*n*=28), and also accounting for baseline DSQ scores, an analogous model revealed no significant effects of DSQ change on either anxiety metric (*Fs*≤1.94, *ps*≥.178). To further investigate the observed relationship between pruning and changes in craving, we checked if any values were considered outliers. After applying an iterative Grubbs approach for outlier removal with a strict threshold of *p<*.05 (5), only one value (DSQ change=61) was identified as a significant outlier (*p=*.033). After removing this datapoint and re-running the above models as well as our previous model predicting **AP** (see main text), results were qualitatively unchanged.

In an LME predicting **LL-discounting** using DAST scores and their interaction with resistance condition as predictors, there was a significant positive effect of DAST (*F*(1,36)=12.40, *p=*.001, 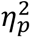=.26; b=-.041), such that greater drug use severity predicted a lower discounting probability through large-loss paths. However, DAST was not found to be predictive of either **NLL-discounting** or **RS**. The subscales of the MAWQ were also largely non-predictive of model parameters (*Fs*≤1.55, *ps*≥.222), apart from the MAWQ functional symptoms subscale, which showed a significant interaction with resistance condition when predicting **NLL-discounting** (*F*(1,38)=4.22, *p=*.047, 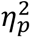=.10). Post-hoc contrasts revealed a negative relationship between functional withdrawal symptoms and **NLL-discounting** when there was an added breathing resistance (ET=-.005; *t*(38)=2.06, *p=*.047). Finally, concerning models predicting changes in DSQ, while accounting for baseline DSQ scores, changes in craving were positively predictive of **LL-discounting** (*F*(1,23)=6.15, *p=*.021, 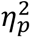=.21; b=.004).

##### Reflectiveness

In analogous tests examining potential relationships between CRT and other model parameters, CRT showed a negative effect on both **LL-discounting** (*F*(1,84)=17.60, *p<*.001, 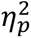=.17) and a positive effect on **RS** (*F*(1,84)=12.38, *p<*.001, 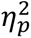=.13), but was not predictive of **NLL-discounting** (*p=*.698). In the clinical group alone, similar results were seen for **LL-discounting** (*p=*.013) and **NLL-discounting** (*p=*.422), while there was no longer a significant relationship with **RS** (*p=*.334). After accounting for working memory and post-test accuracy, there was still a significant negative effect of CRT scores on **AP** across both groups (*F*(1,73)=12.06, *p<*.001, 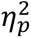=.14), but was only marginal in iMUDs alone (*F*(1,28)=4.05, *p=*.054, 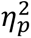=.13). In both cases, these relationships were driven by **LL-discounting**. The previously observed relationship between CRT and **RS** across all participants was also retained after accounting for differences in working memory (*p=*.004).

#### Exploratory analyses of impulsivity and reward-seeking

To test the hypothesis that individuals with higher impulsivity would show greater aversive pruning, we performed a latent factor analysis including the subscales from the UPPS-P, BIS/BAS, and TEPS (using the *fa* function from the psych package in R; (6)). The number of factors was chosen using the *fa.parallel* function (fm=“pa”, fa=“fa”, n.iter=1000) and Bartlett factor scores (7) were calculated using an oblimin rotation allowing factors to be correlated. Then, factor scores were included in LMEs predicting model parameters in the clinical group alone to assess the impact of impulsive tendencies on task behavior. The measure of sampling adequacy (MSA=.65) demonstrated mediocre suitability for factor analysis and principal analysis suggested 4 factors (see **Table S7**; (8)). However, none of the 4 identified factors predicted any model parameter in iMUDs (*Fs*≤2.52, *ps*≥.122).

### Supplemental Figures

**Fig S1.**
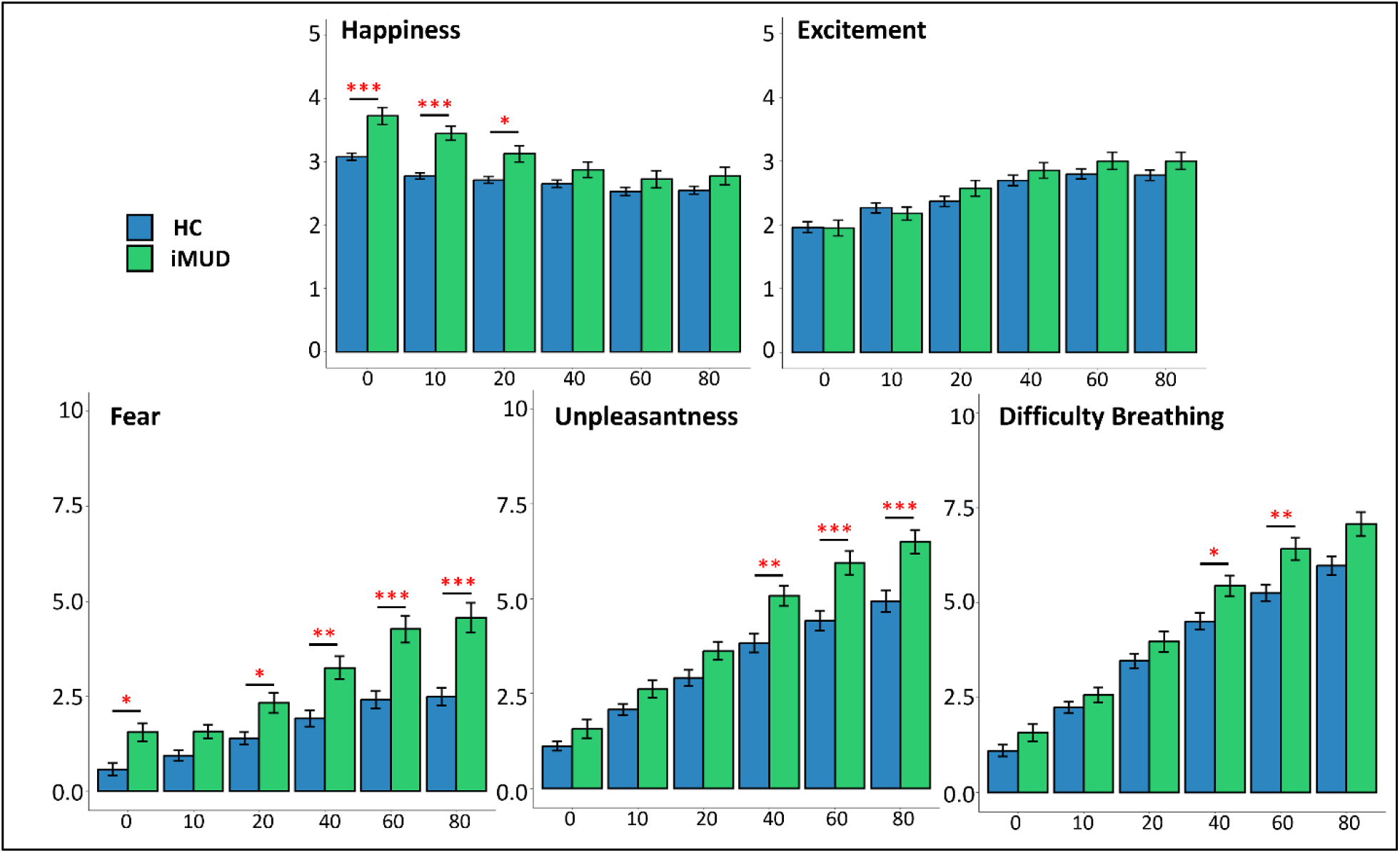
Additional affective ratings from resistance sensitivity protocol. Means and standard error bars for self-reported anxiety ratings across the resistance sensitivity protocol (units are cmH20/L/sec). Note that the scales for *unpleasantness, difficulty breathing,* and *fear* are 0-10, while those for *happiness* and *excitement* are 1-5. Stars indicate significant group differences at each resistance level.

**Fig S2.**
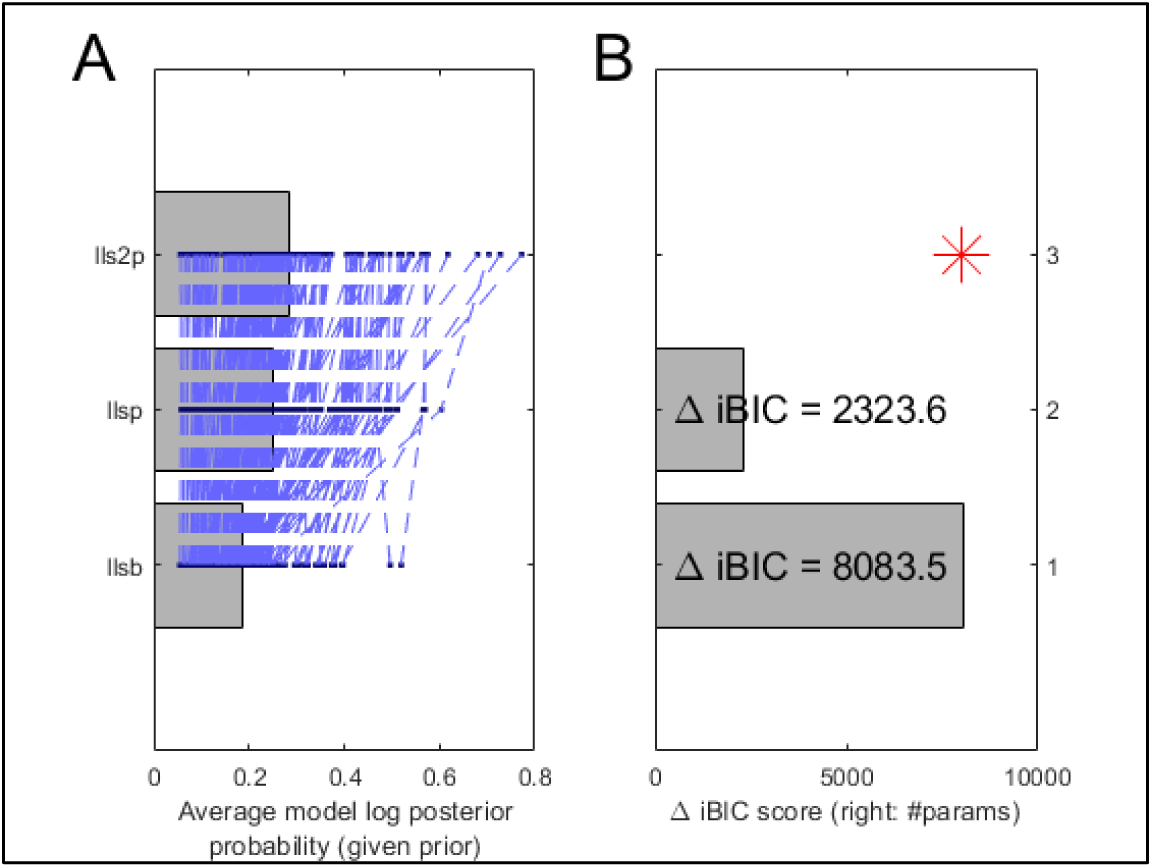
Model comparison. **A)** Comparison of log posterior probabilities for each model tested. **B)** BIC comparison between the winning model (“pruning” model: top) and the two other models tested (“discounting” model: middle; “lookahead” model: bottom).

**Fig S3.**
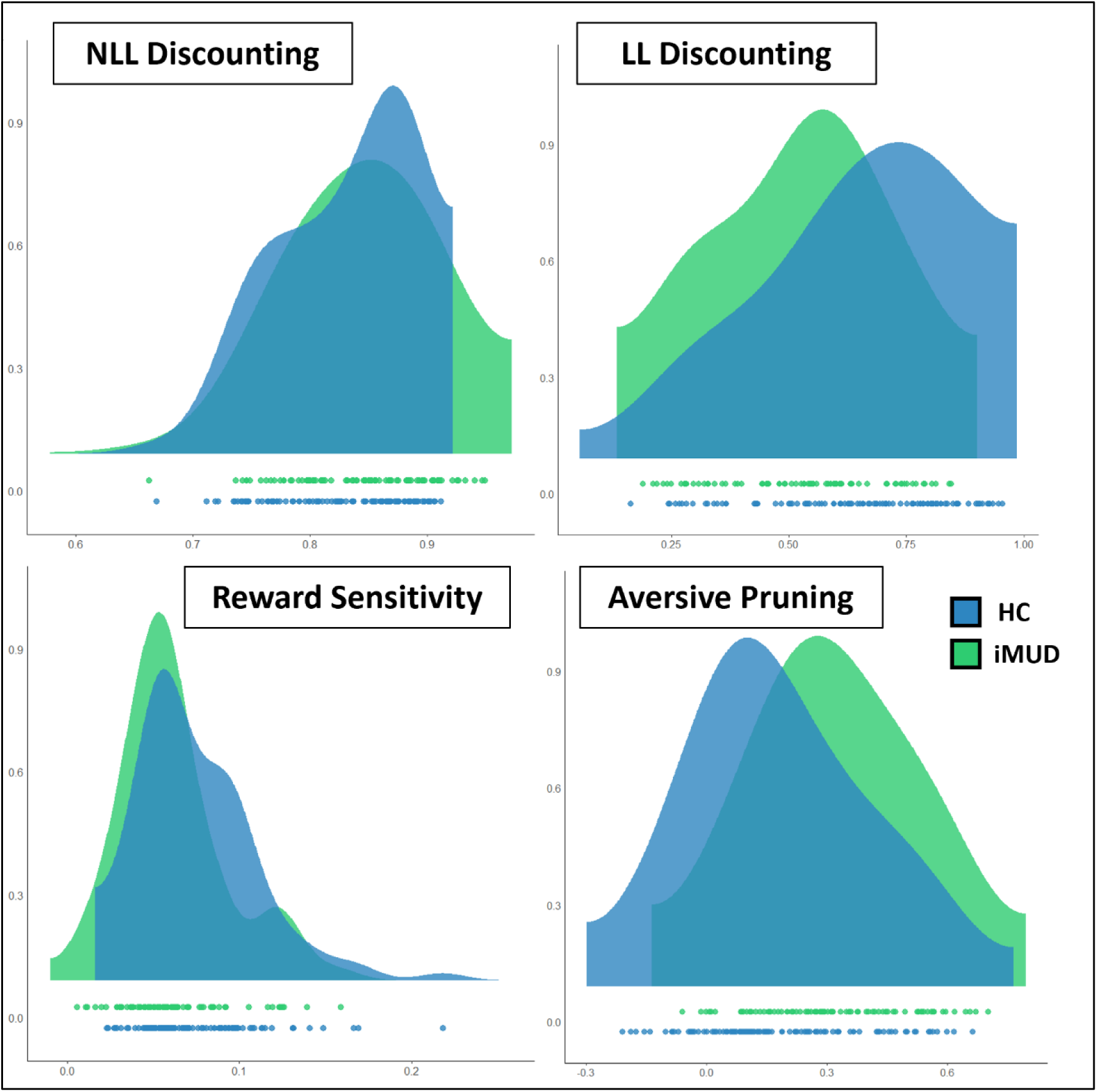
Computational parameter distributions. Density plots of model parameters colored by group classification with points at the bottom indicating individual values. Note that these distributions are collapsed across resistance conditions.

**Figure S4.**
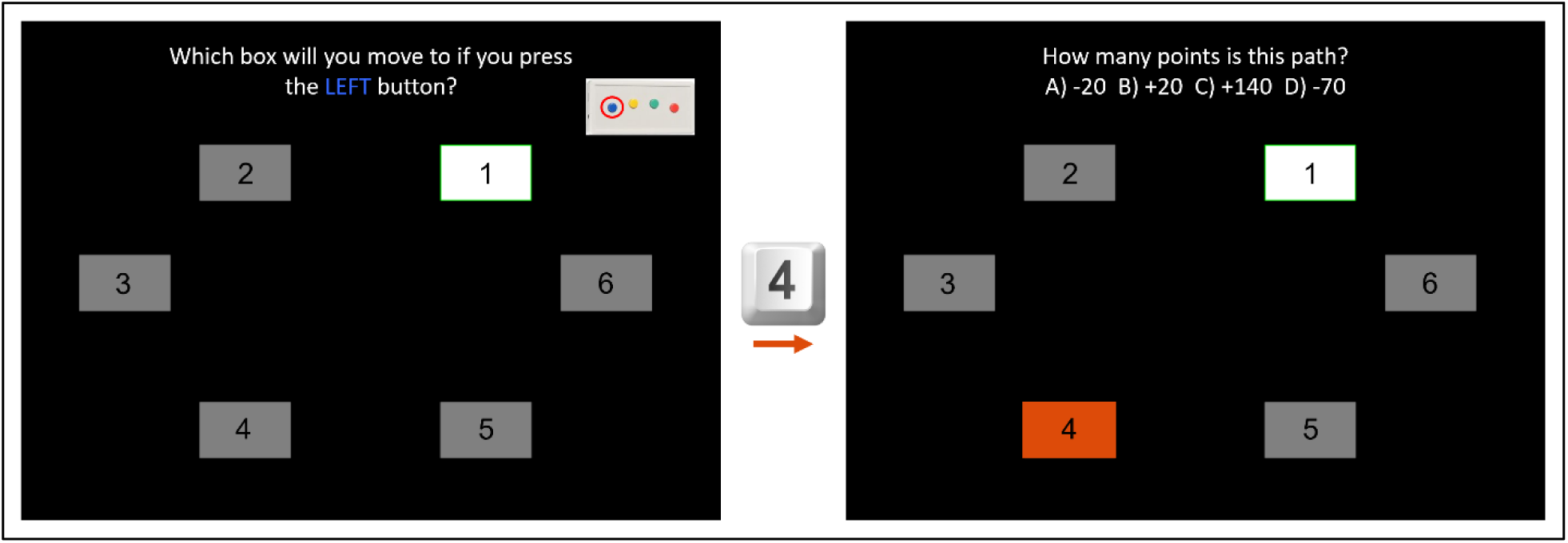
Post-test assessment. An example question from the post-test assessment. For each starting position and direction, participants are asked to identify the transition and the associated points (24 questions). Because the question about points is dependent on which transition the participant believed they would make, there is then a portion of the post-test which goes through each possible transition and asks how many points are awarded on that path (12 questions). Overall accuracy on this test was determined by the percentage of correct answers on the entire assessment. The derived accuracy metric is instead the ratio of accuracy on the questions pertaining to −70 paths (both transitions and points) to accuracy on all other questions. That way, a derived accuracy of <1 would mean worse memory of large loss paths compared to all other paths.

**Fig. S5.**
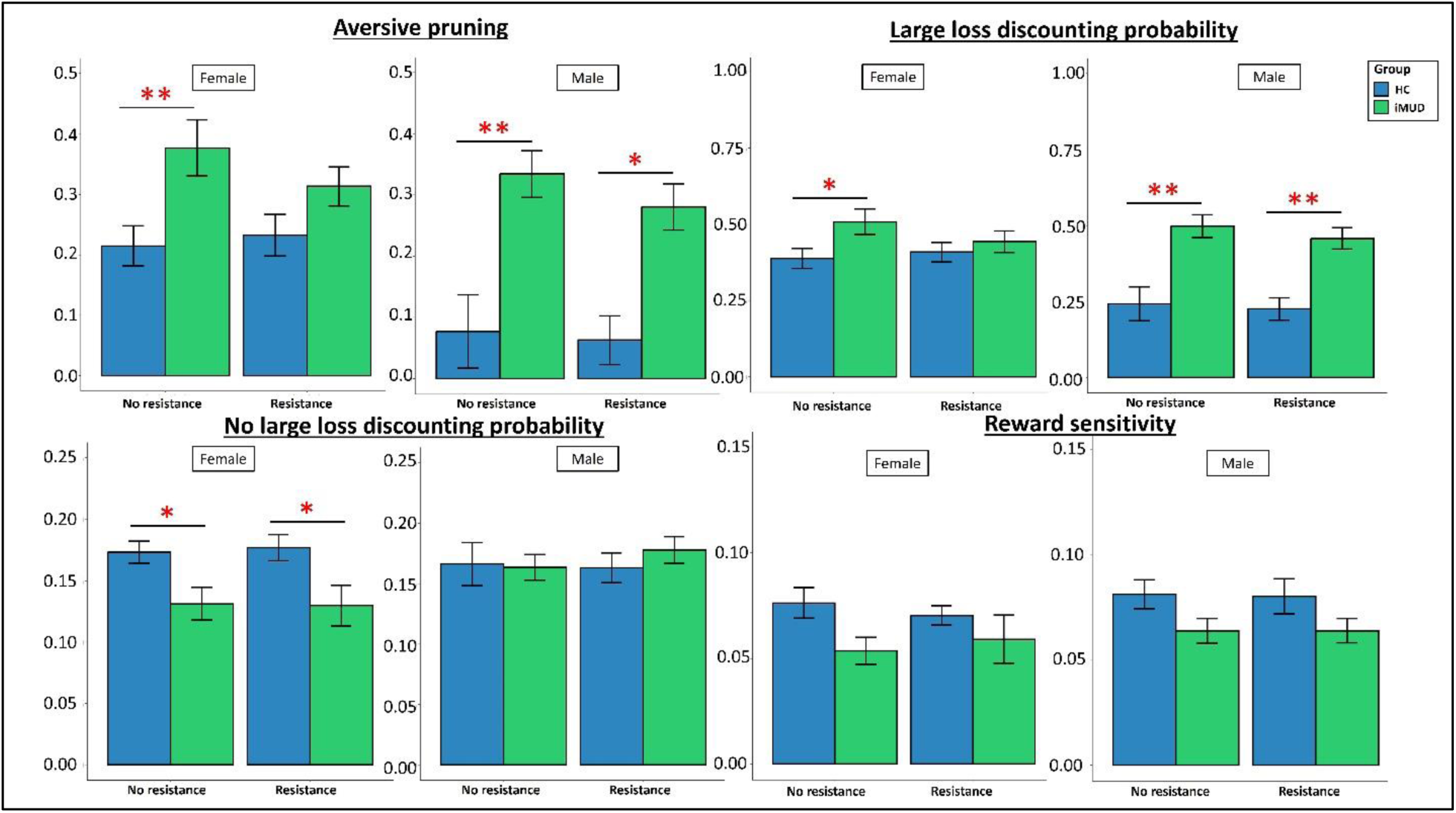
Computational parameters by sex. Means and standard errors for each model parameter separated by both group and sex. Stars indicate significant post-hoc group differences in each resistance condition from models that predicted each parameter after subsetting participants by sex (accounting for age and state anxiety). *p<.05, **p<.01, ***p<.001.

**Fig. S6.**
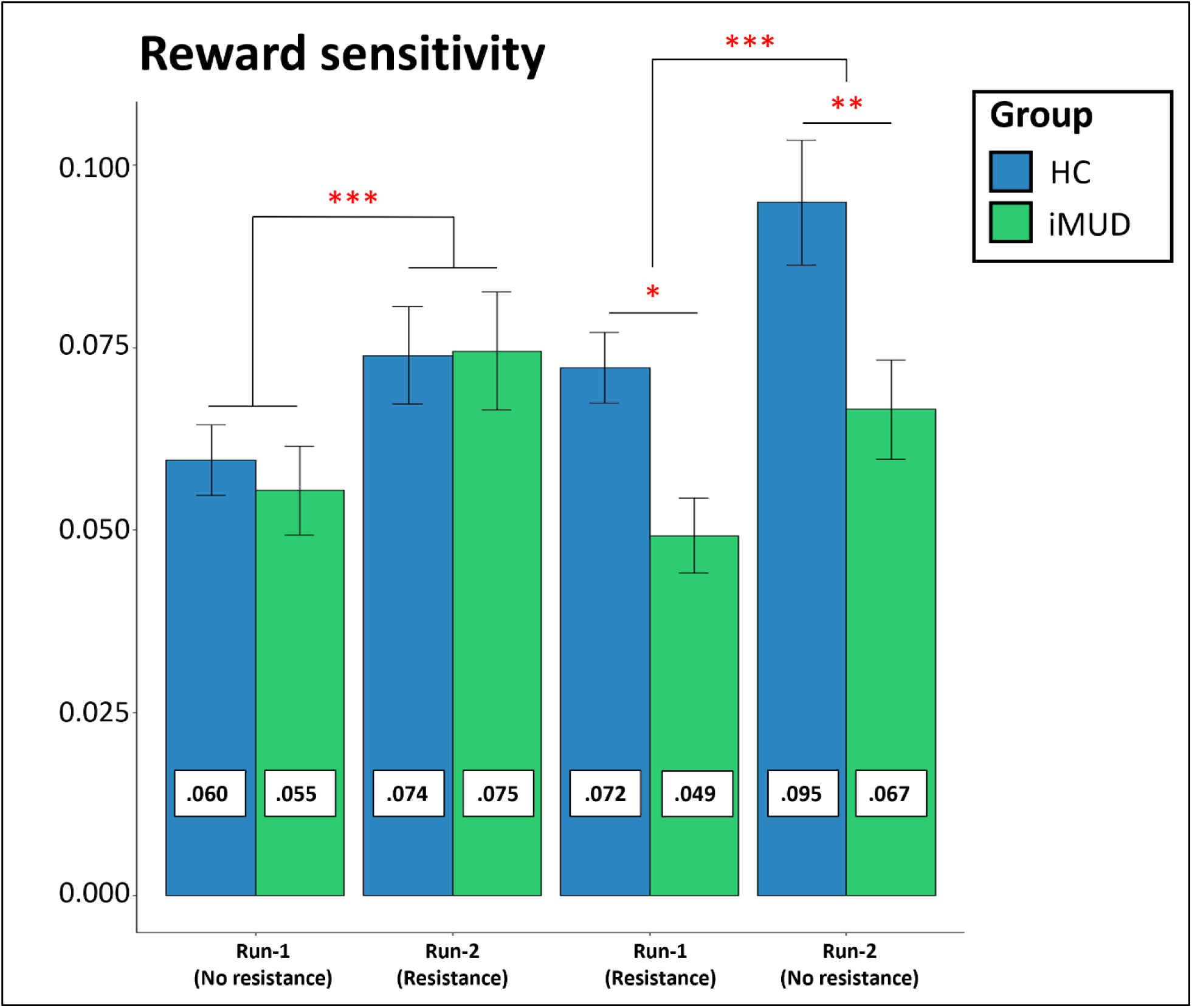
Reward sensitivity by condition order. Means and standard errors for **RS** values by group, condition order, and resistance condition. Estimated marginal means (in boxes on each bar) come from post-hoc contrasts following an LME predicting **RS** estimates with *group x order x resistance*. Stars indicate significant differences in these post-hoc contrasts.

### Supplemental Tables

**Table S1.** Results of models predicting each affective question during the breathing resistance protocol.

| **Affect** | **Predictor** | **F** | **Post-hoc Test** |
| --- | --- | --- | --- |
| **Unpleasantness** | Group | $F(1,87)=6.16, p=.015$ | iMUD (EMM=4.22),<br>HC (EMM=3.22);<br>iMUD - HC:<br>$t(87)=2.48, p=.015$ |
| | Resistance | $F(1,977)=1389.63, p<.001$ | b=0.901 |
| | Group*Resistance | $F(1,977)=28.70, p<.001$ | iMUD (ET=1.03), HC (ET=0.77);<br>iMUD - HC:<br>$t(977)=5.37, p<.001$ |
| **Difficulty** | Group | $F(1,87)=3.97, p=.049$ | iMUD (EMM=4.51),<br>HC (EMM=3.76);<br>iMUD - HC:<br>$t(87)=1.99, p=.049$ |
| | Resistance | $F(1,977)=2040.89, p<.001$ | b=1.072 |
| | Group*Resistance | $F(1,977)=13.29, p<.001$ | iMUD (ET=1.16), HC (ET=0.99);<br>iMUD - HC:<br>$t(977)=3.65, p<.001$ |
| **Fear** | Group | $F(1,87)=11.04, p=.001$ | iMUD (EMM=2.92),<br>HC (EMM=1.62);<br>iMUD - HC:<br>$t(87)=3.32, p=.001$ |
| | Resistance | $F(1,977)=450.45, p<.001$ | b=0.553 |
| | Group*Resistance | $F(1,977)=29.01, p<.001$ | iMUD (ET=0.69), HC (ET=0.42);<br>iMUD - HC:<br>$t(977)=5.39, p<.001$ |
| **Happiness** | Group | $F(1,87)=7.23, p=.009$ | iMUD (EMM=3.11),<br>HC (EMM=2.72);<br>iMUD - HC:<br>$t(87)=2.69, p=.009$ |
| | Resistance | $F(1,977)=189.93, p<.001$ | b=-0.152 |
| | Group*Resistance | $F(1,977)=26.03, p<.001$ | iMUD (ET=-0.21), HC (ET=-0.10); |
| | | | iMUD - HC:<br>$t(977)=5.10, p<.001$ |
| **Excitement** | Group | $F(1,87)=0.54, p=.462$ | ns |
| | Resistance | $F(1,977)=286.68, p<.001$ | b=0.200 |
| | Group*Resistance | $F(1,977)=5.96, p=.015$ | iMUD (ET=0.23), HC<br>(ET=0.17); |
| | | | iMUD - HC:<br>$t(977)=2.44, p=.015$ |

**Table S2A.**
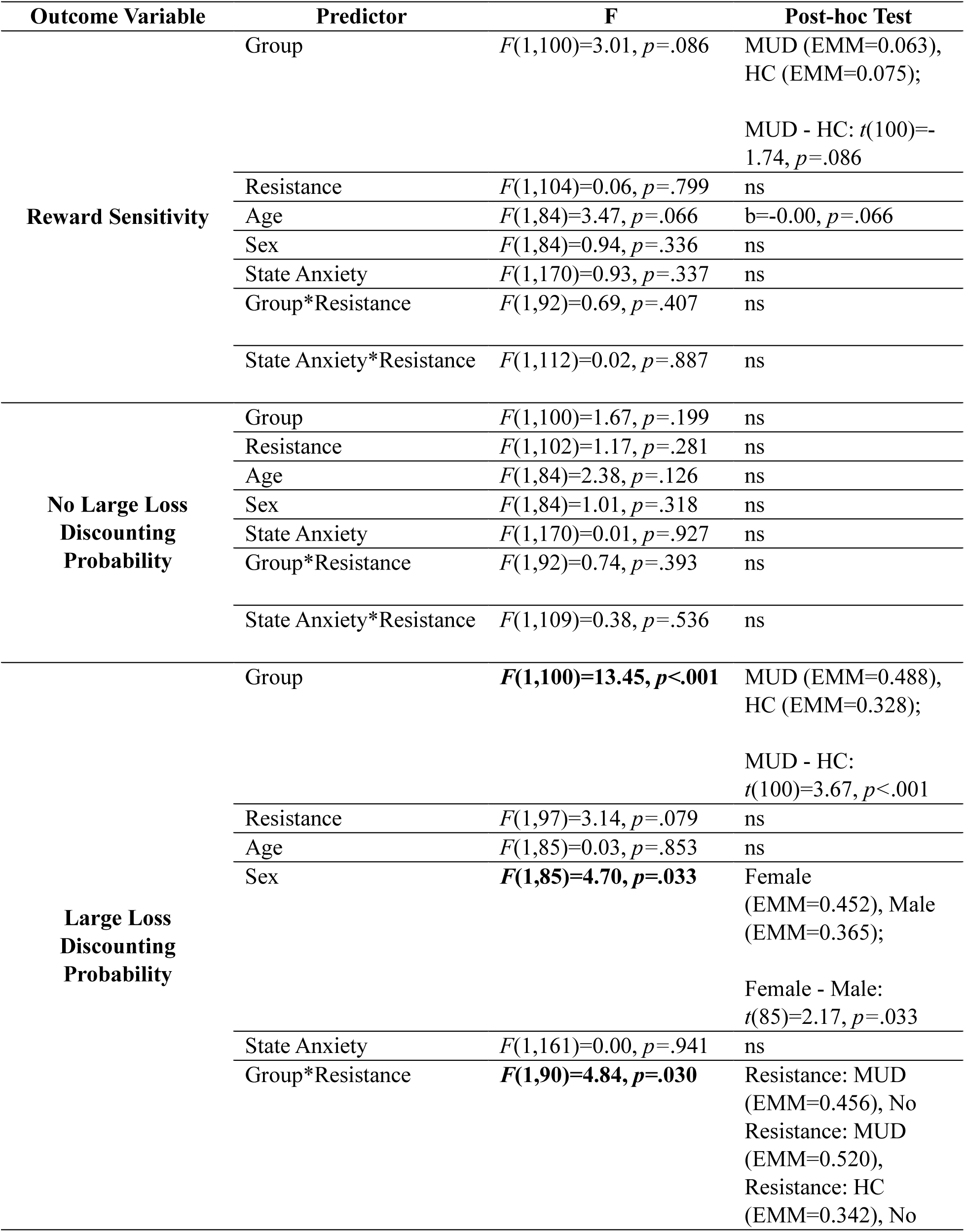
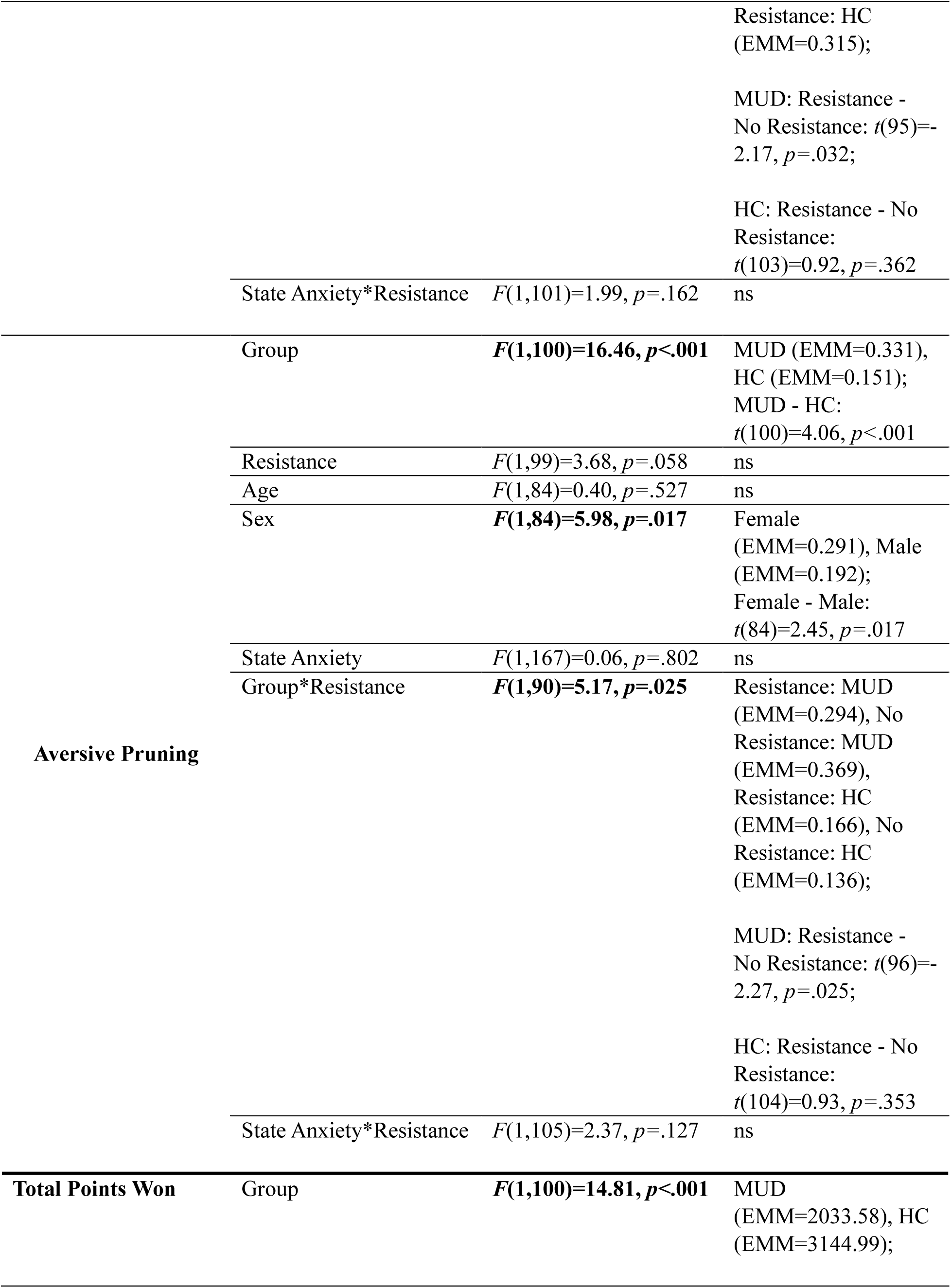
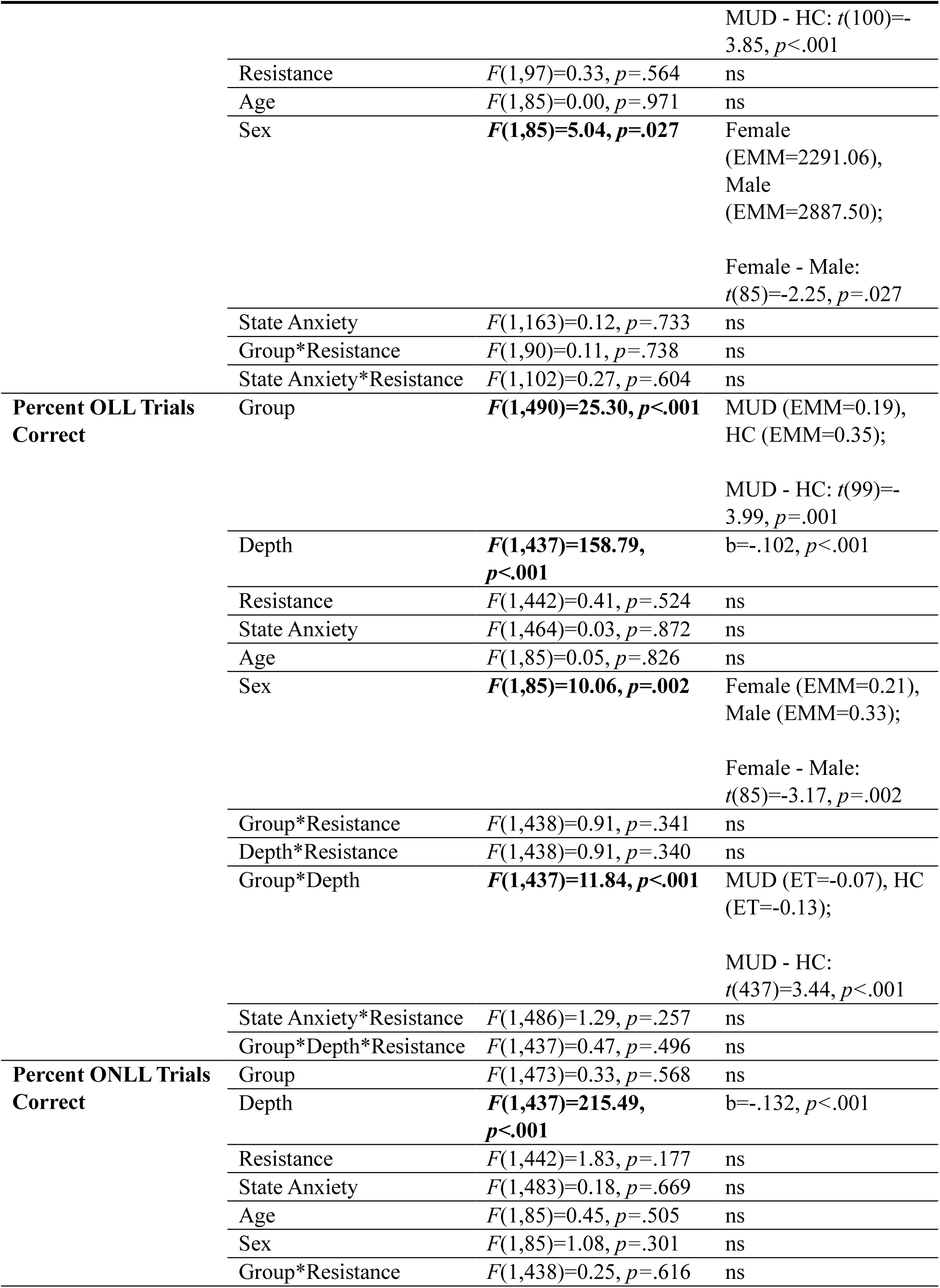
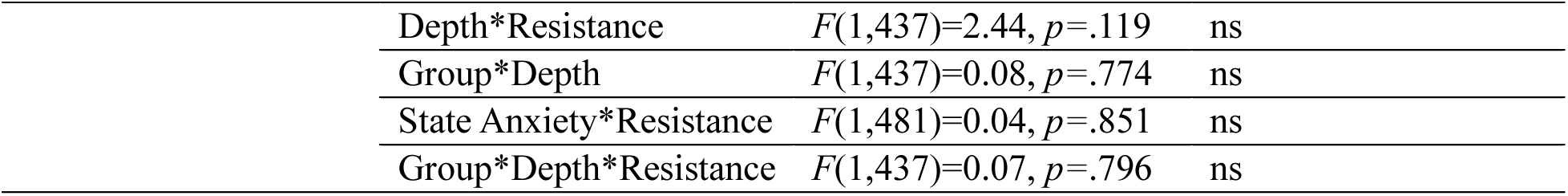
Full results of the models from Table 3 in the main text.

**Table S2B.**
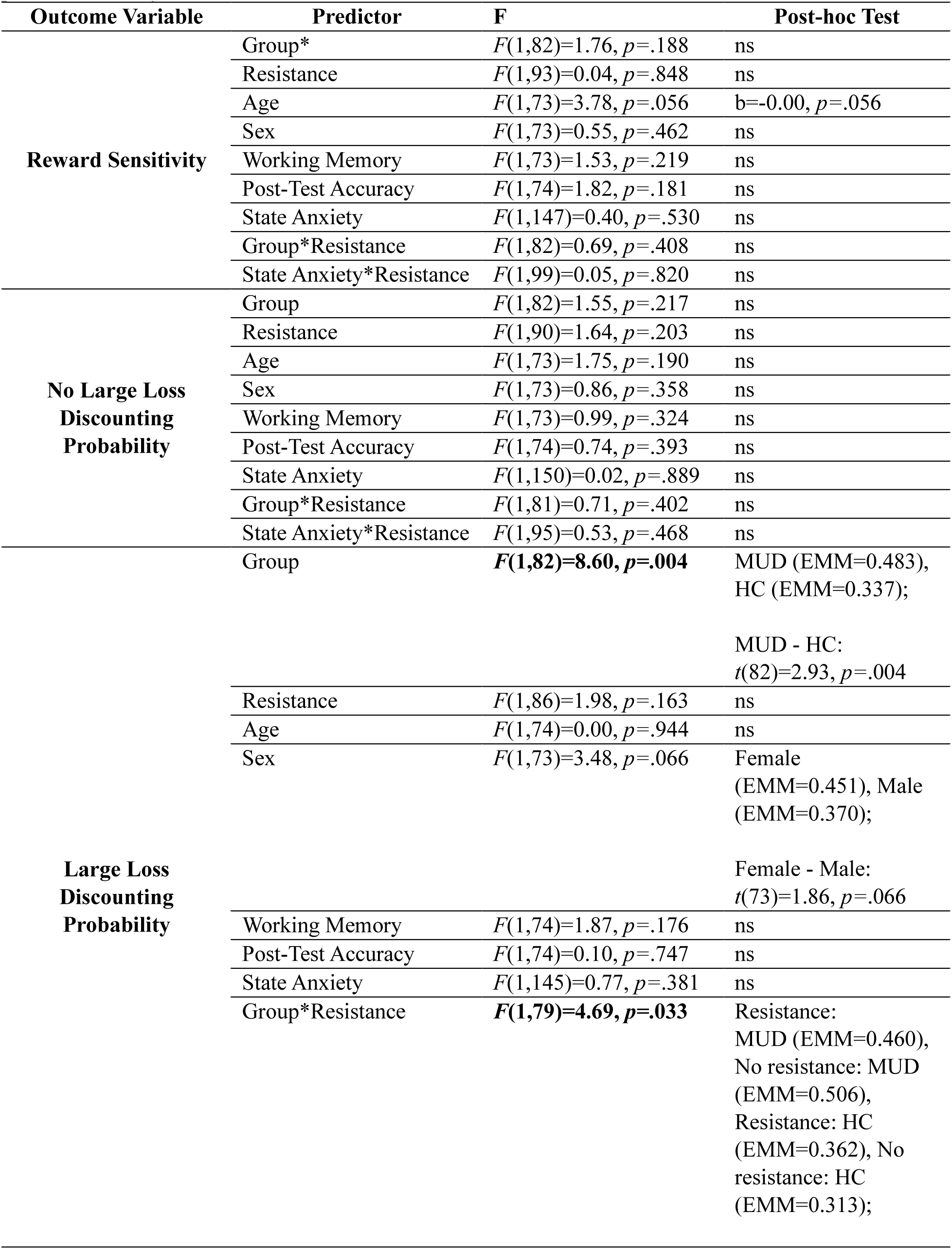
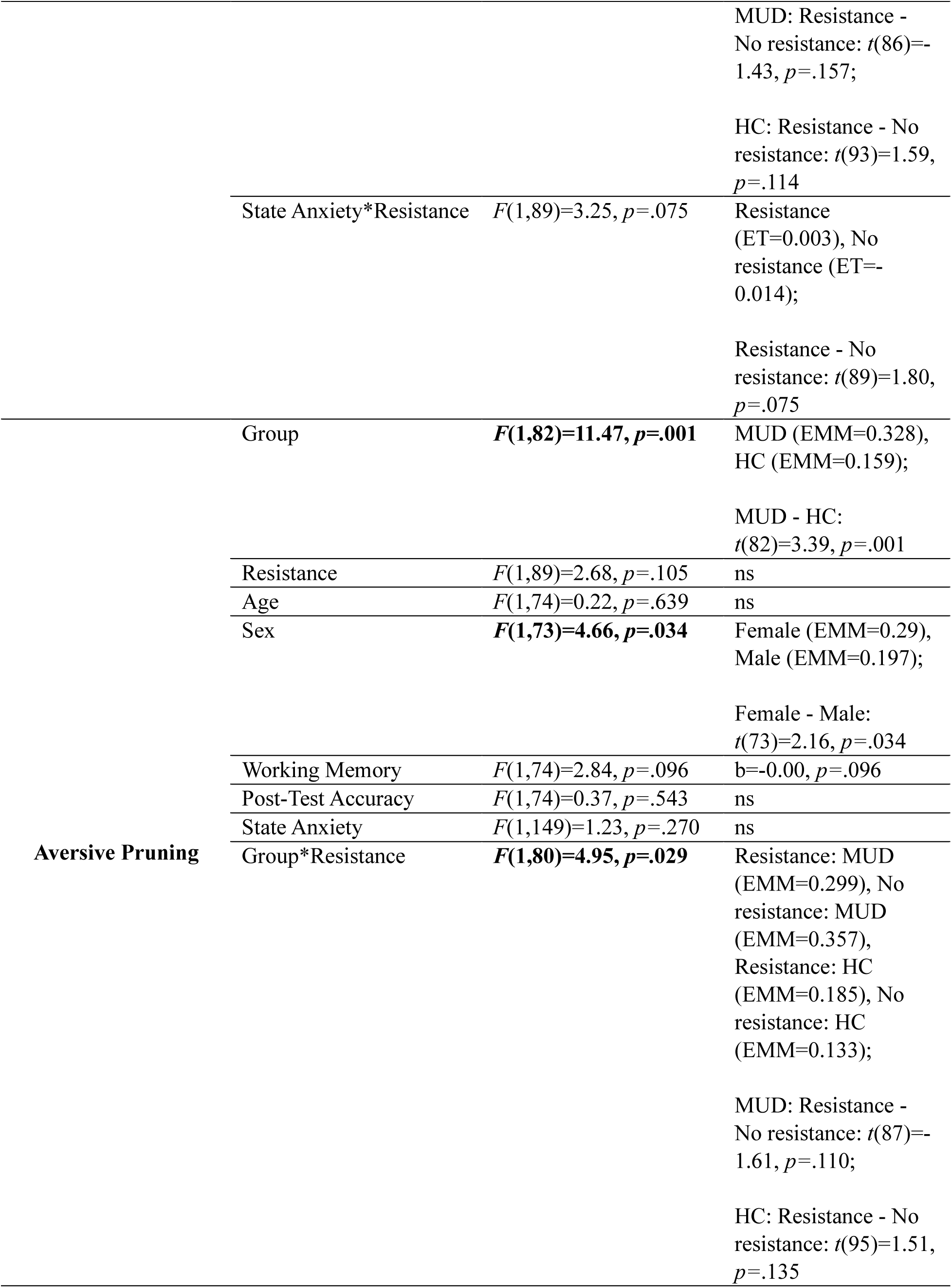
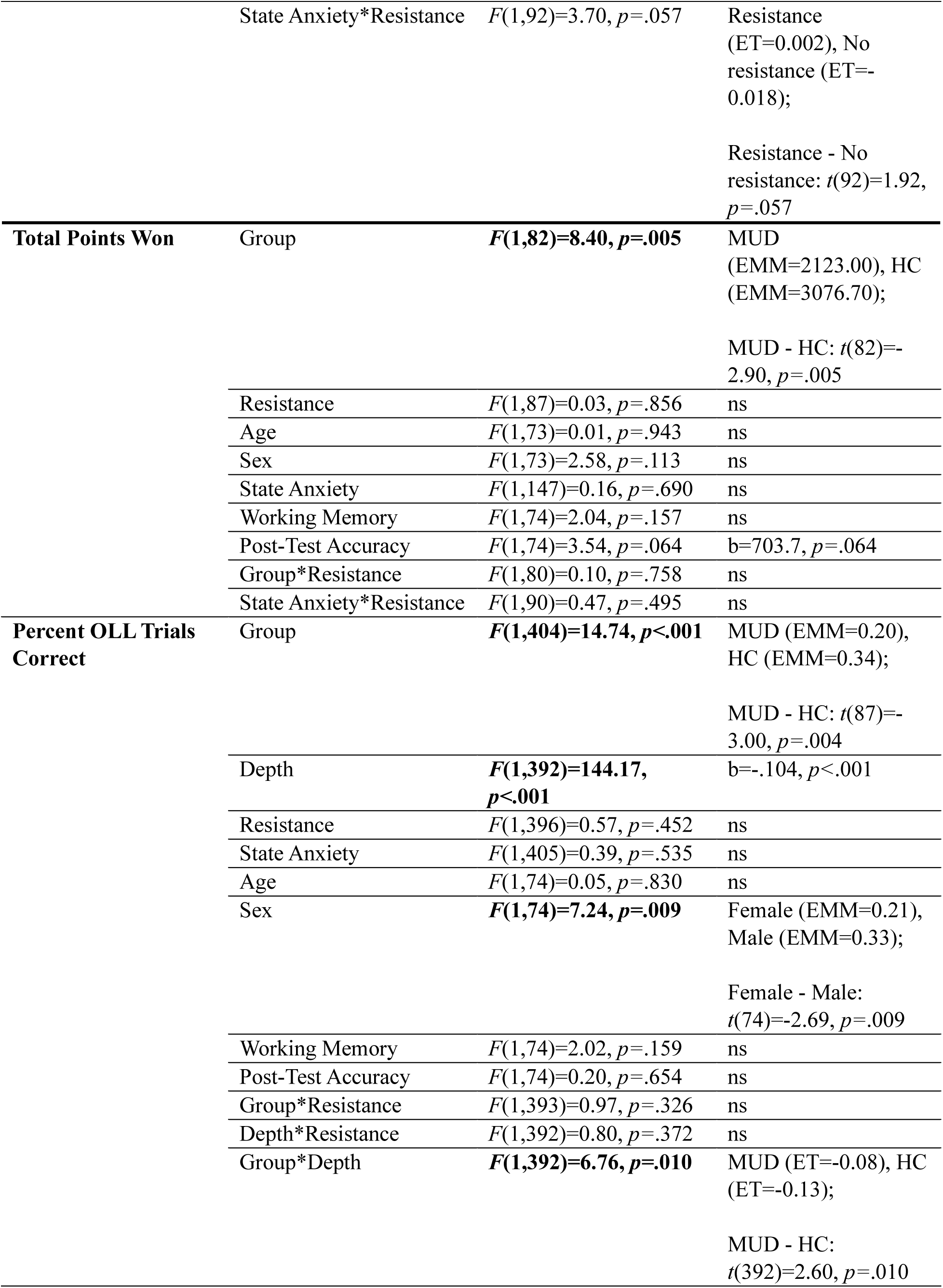
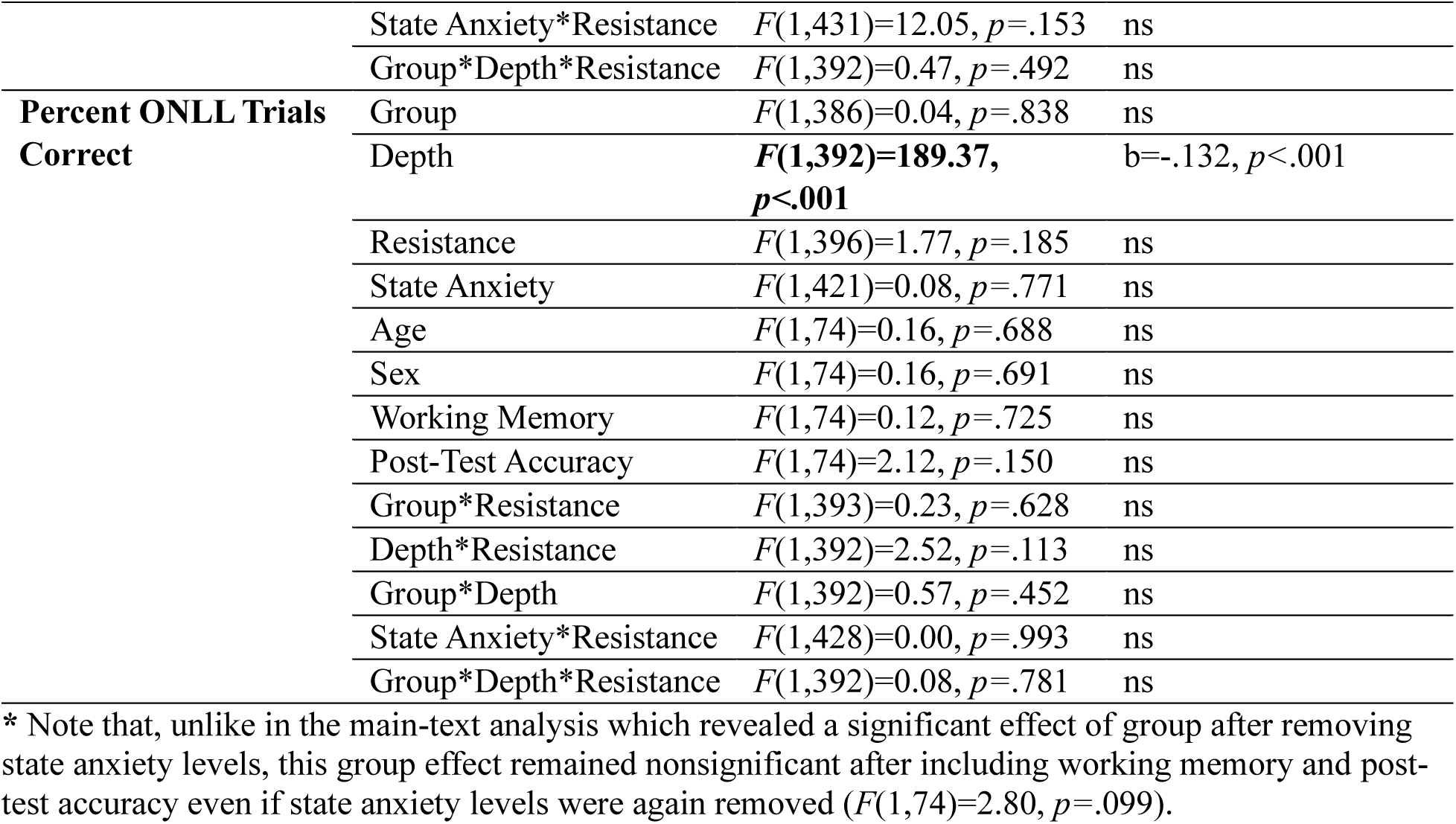
Full results of the models from Table 3 in the main text after including working memory and post-test accuracy.

**Table S3A.**
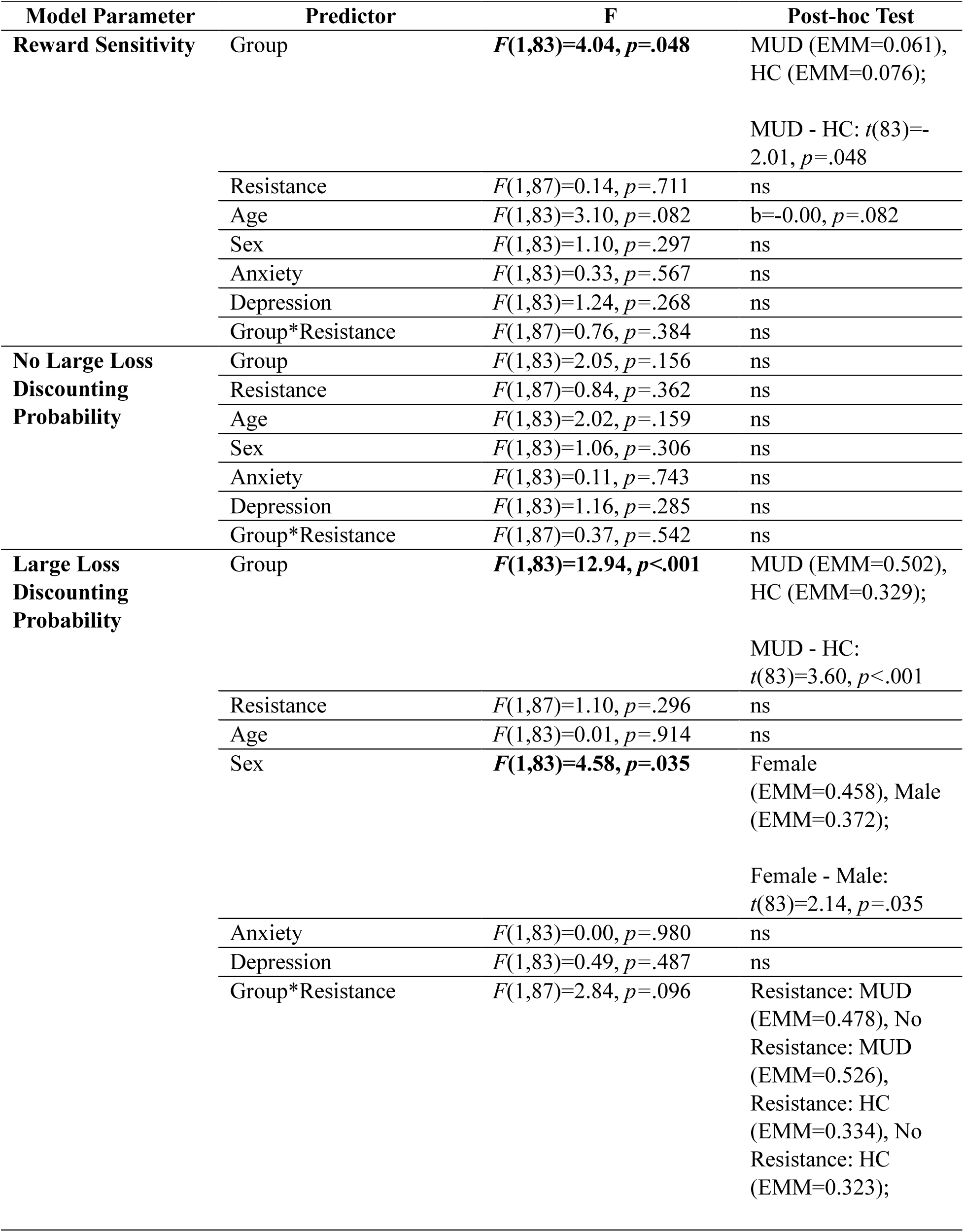
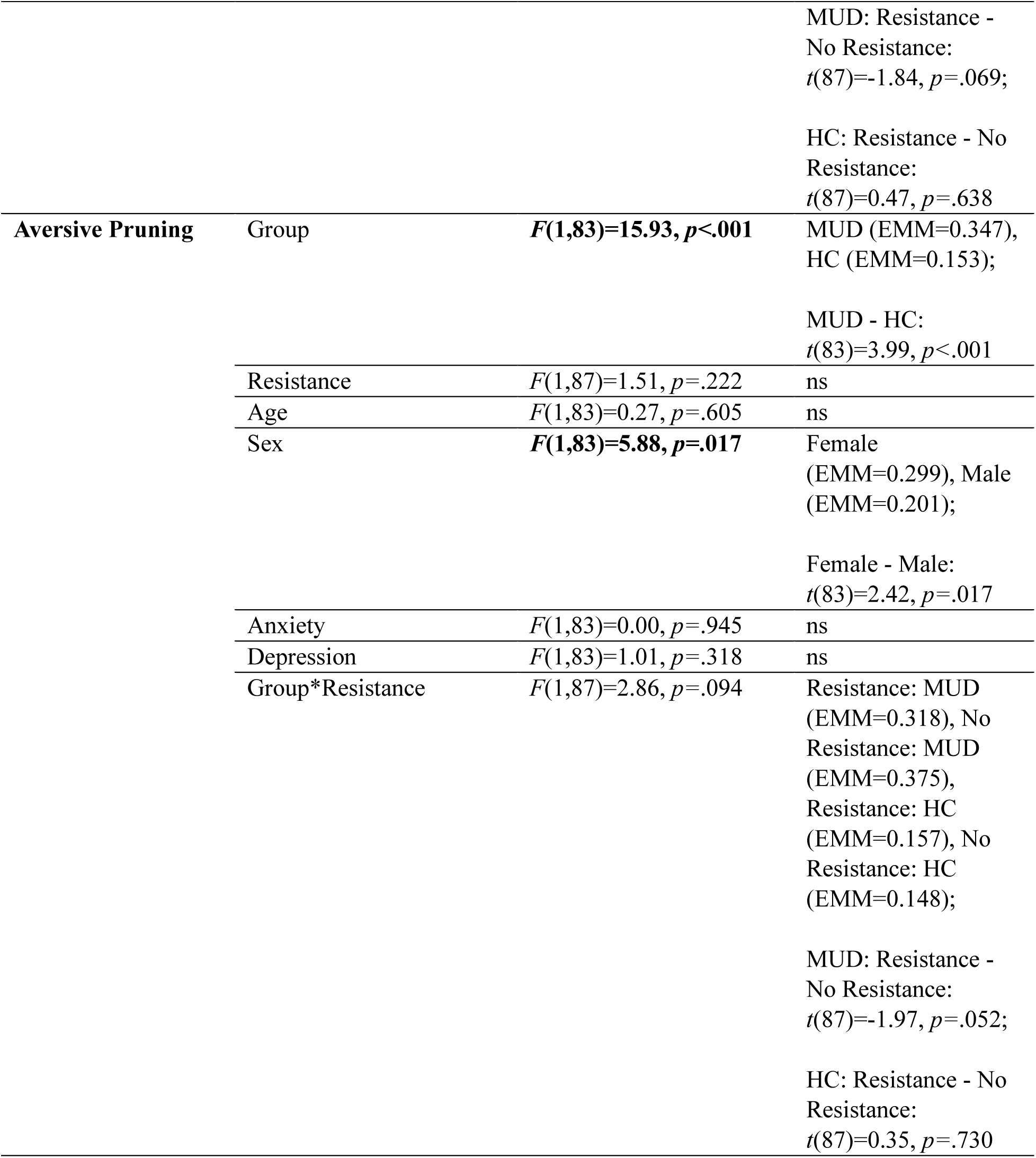
Full results of models including trait clinical symptoms for each computational parameter.

| Model Parameter | Predictor | F | Post-hoc Test |
| --- | --- | --- | --- |
| Reward Sensitivity | Group | $F(1,83)=4.04, p=.048$ | MUD (EMM=0.061),<br>HC (EMM=0.076);<br>MUD - HC: $t(83)=-2.01, p=.048$ |
| | Resistance | $F(1,87)=0.14, p=.711$ | ns |
| | Age | $F(1,83)=3.10, p=.082$ | $b=-0.00, p=.082$ |
| | Sex | $F(1,83)=1.10, p=.297$ | ns |
| | Anxiety | $F(1,83)=0.33, p=.567$ | ns |
| | Depression | $F(1,83)=1.24, p=.268$ | ns |
| | Group*Resistance | $F(1,87)=0.76, p=.384$ | ns |
| No Large Loss Discounting Probability | Group | $F(1,83)=2.05, p=.156$ | ns |
| | Resistance | $F(1,87)=0.84, p=.362$ | ns |
| | Age | $F(1,83)=2.02, p=.159$ | ns |
| | Sex | $F(1,83)=1.06, p=.306$ | ns |
| | Anxiety | $F(1,83)=0.11, p=.743$ | ns |
| | Depression | $F(1,83)=1.16, p=.285$ | ns |
| | Group*Resistance | $F(1,87)=0.37, p=.542$ | ns |
| Large Loss Discounting Probability | Group | $F(1,83)=12.94, p<.001$ | MUD (EMM=0.502),<br>HC (EMM=0.329);<br>MUD - HC:<br>$t(83)=3.60, p<.001$ |
| | Resistance | $F(1,87)=1.10, p=.296$ | ns |
| | Age | $F(1,83)=0.01, p=.914$ | ns |
| | Sex | $F(1,83)=4.58, p=.035$ | Female<br>(EMM=0.458), Male<br>(EMM=0.372);<br>Female - Male:<br>$t(83)=2.14, p=.035$ |
| | Anxiety | $F(1,83)=0.00, p=.980$ | ns |
| | Depression | $F(1,83)=0.49, p=.487$ | ns |
| | Group*Resistance | $F(1,87)=2.84, p=.096$ | Resistance: MUD<br>(EMM=0.478), No<br>Resistance: MUD<br>(EMM=0.526),<br>Resistance: HC<br>(EMM=0.334), No<br>Resistance: HC<br>(EMM=0.323); |
| | | | MUD: Resistance - No Resistance:<br>$t(87)=-1.84, p=.069$; |
| | | | HC: Resistance - No Resistance:<br>$t(87)=0.47, p=.638$ |
| Aversive Pruning | Group | **$F(1,83)=15.93, p<.001$** | MUD (EMM=0.347), HC (EMM=0.153); |
| | | | MUD - HC:<br>$t(83)=3.99, p<.001$ |
| | Resistance | $F(1,87)=1.51, p=.222$ | ns |
| | Age | $F(1,83)=0.27, p=.605$ | ns |
|  | Sex | **$F(1,83)=5.88, p=.017$** | Female (EMM=0.299), Male (EMM=0.201); |
| | | | Female - Male:<br>$t(83)=2.42, p=.017$ |
| | Anxiety | $F(1,83)=0.00, p=.945$ | ns |
| | Depression | $F(1,83)=1.01, p=.318$ | ns |
| | Group*Resistance | $F(1,87)=2.86, p=.094$ | Resistance: MUD (EMM=0.318), No Resistance: MUD (EMM=0.375), Resistance: HC (EMM=0.157), No Resistance: HC (EMM=0.148); |
| | | | MUD: Resistance - No Resistance:<br>$t(87)=-1.97, p=.052$; |
| | | | HC: Resistance - No Resistance:<br>$t(87)=0.35, p=.730$ |

**Table S3B.**
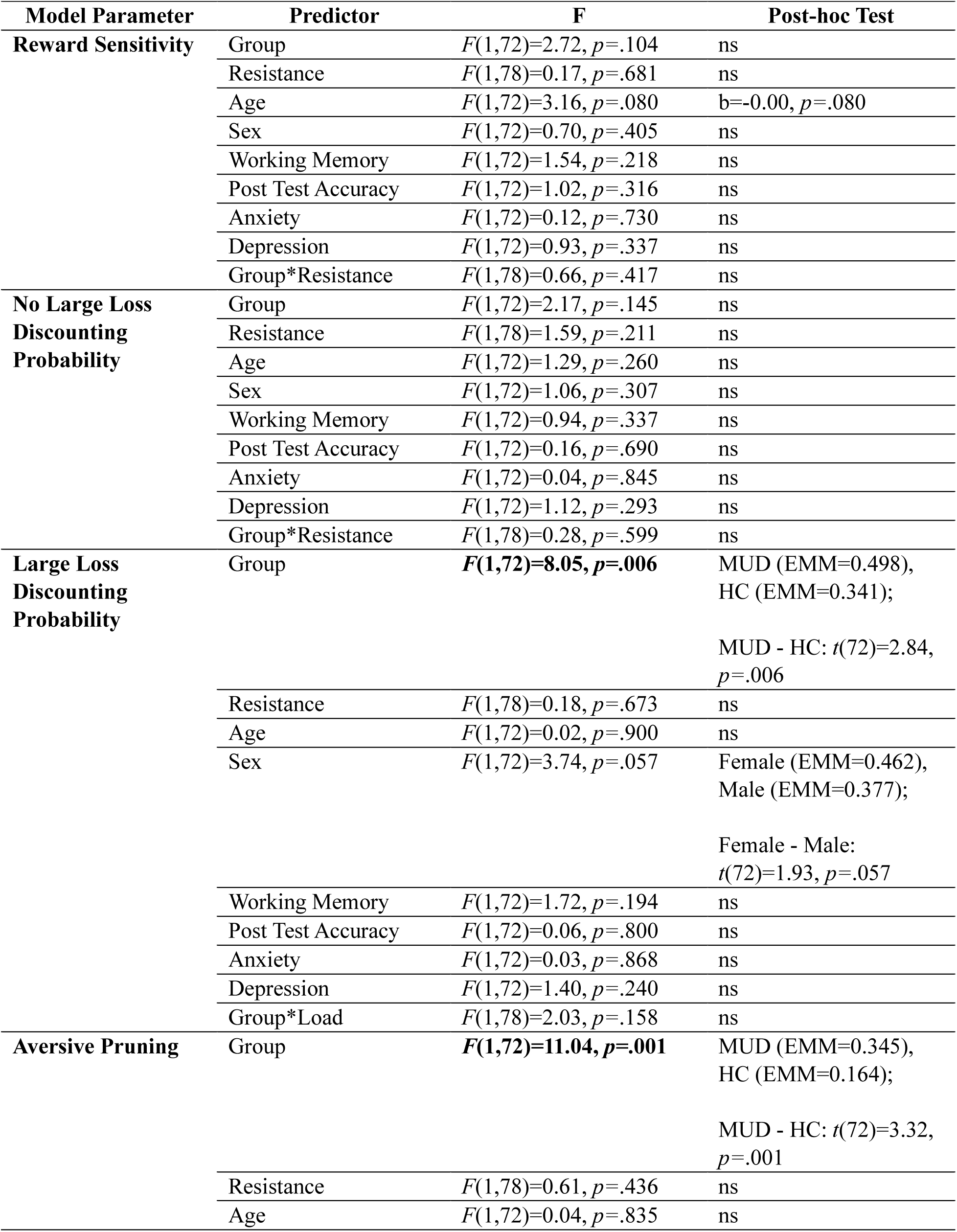
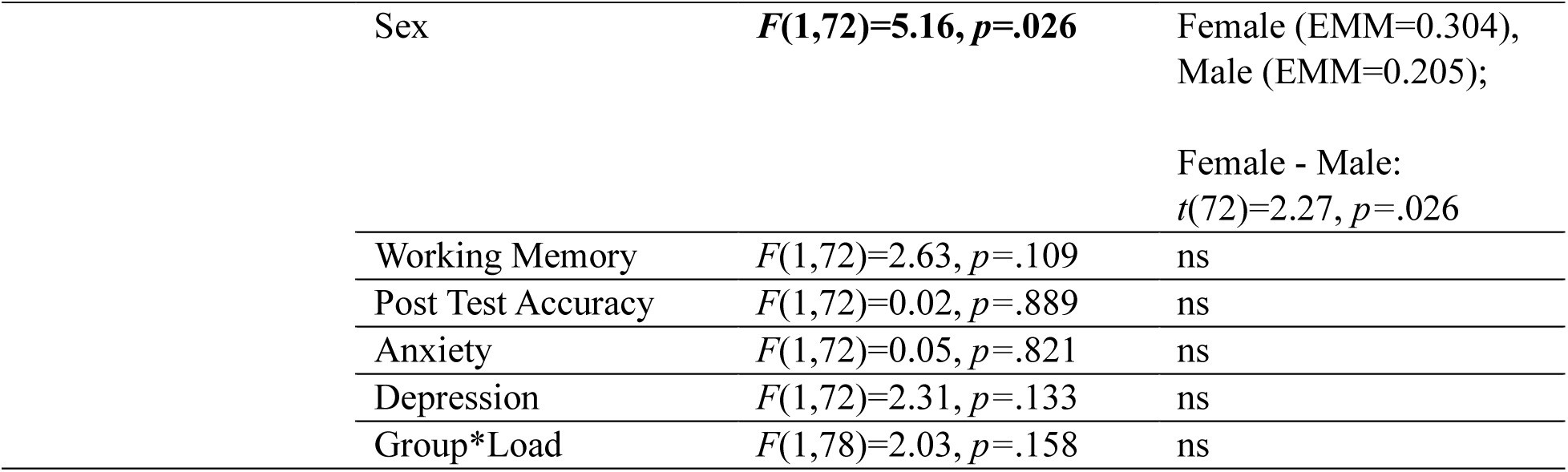
Full results of models including trait clinical symptoms for each computational parameter (along with working memory and post-test accuracy).

**Table S4.** Comorbidity table for participants with MUD (n=40).

|  | **Alcohol** | **Cannabis** | **Opioid** | **Sedative** | **Cocaine** | **MDD** | **GAD** | **SAD** | **Panic** | **PTSD** |
| --- | --- | --- | --- | --- | --- | --- | --- | --- | --- | --- |
| **Alcohol** | **15** |  |  |  |  |  |  |  |  |  |
| **Cannabis** | 4 | **8** |  |  |  |  |  |  |  |  |
| **Opioid** | 4 | 0 | **15** |  |  |  |  |  |  |  |
| **Sedative** | 0 | 0 | 1 | **1** |  |  |  |  |  |  |
| **Cocaine** | 0 | 0 | 1 | 1 | **2** |  |  |  |  |  |
| **MDD** | 5 | 1 | 3 | 0 | 0 | **7** |  |  |  |  |
| **GAD** | 1 | 0 | 1 | 0 | 0 | 1 | **1** |  |  |  |
| **SAD** | 0 | 0 | 0 | 0 | 0 | 0 | 0 | **0** |  |  |
| **Panic** | 0 | 0 | 0 | 0 | 0 | 0 | 0 | 0 | **0** |  |
| **PTSD** | 0 | 0 | 0 | 0 | 0 | 0 | 0 | 0 | 0 | **0** |

**Note.** All participants in this group had a primary diagnosis of amphetamine use disorder with methamphetamine as their drug of choice. The diagnoses here reflect additional comorbidities. The diagonal contains the total number of participants with a given diagnosis (regardless of comorbidities). Alcohol=alcohol use disorder, Cannabis=cannabis use disorder, Opioid=opioid use disorder, Sedative=sedative use disorder, Cocaine=cocaine use disorder, MDD=major depressive disorder, GAD=generalized anxiety disorder, SAD=social anxiety disorder, Panic=panic disorder, PTSD=post-traumatic stress disorder.

**Table S5.**
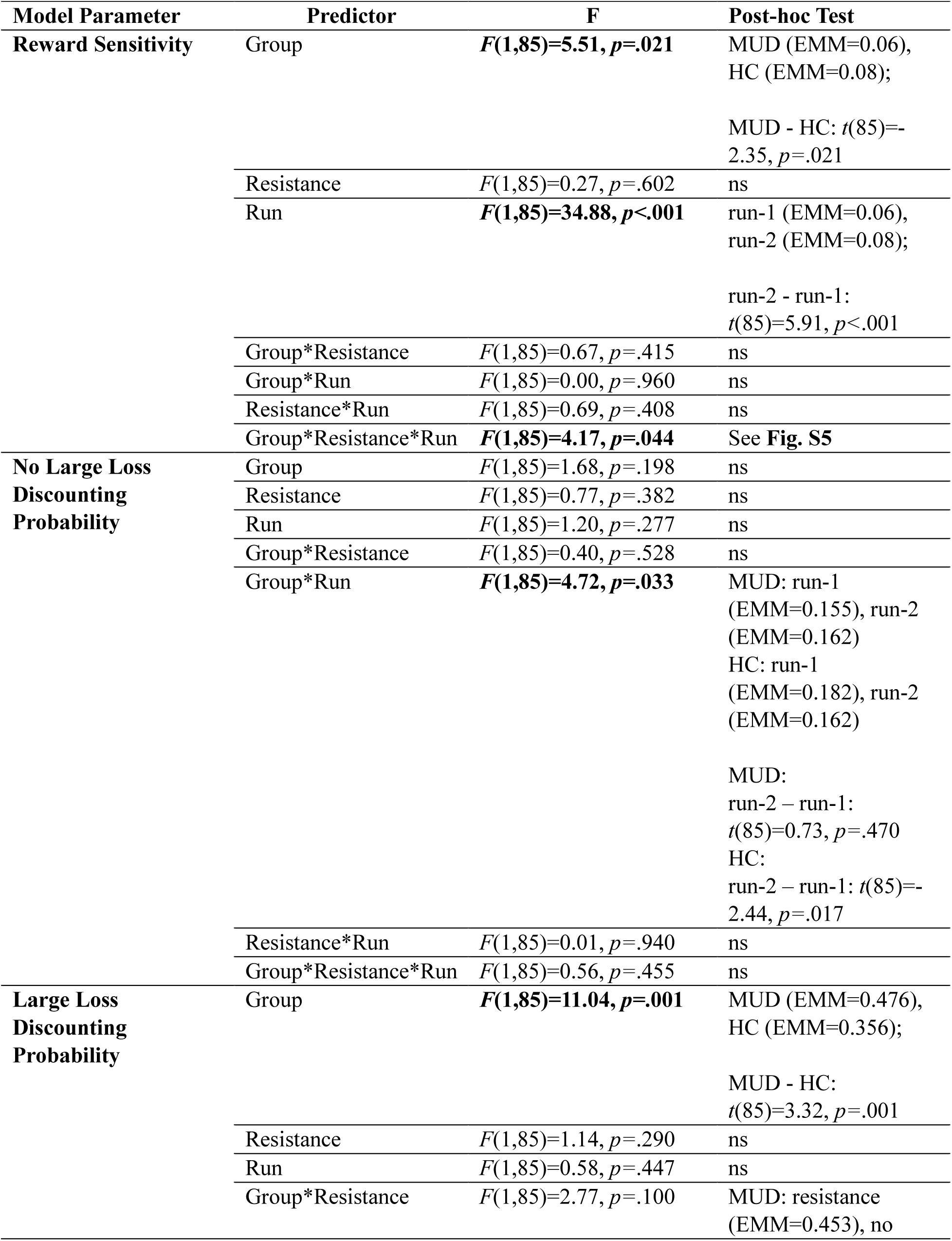
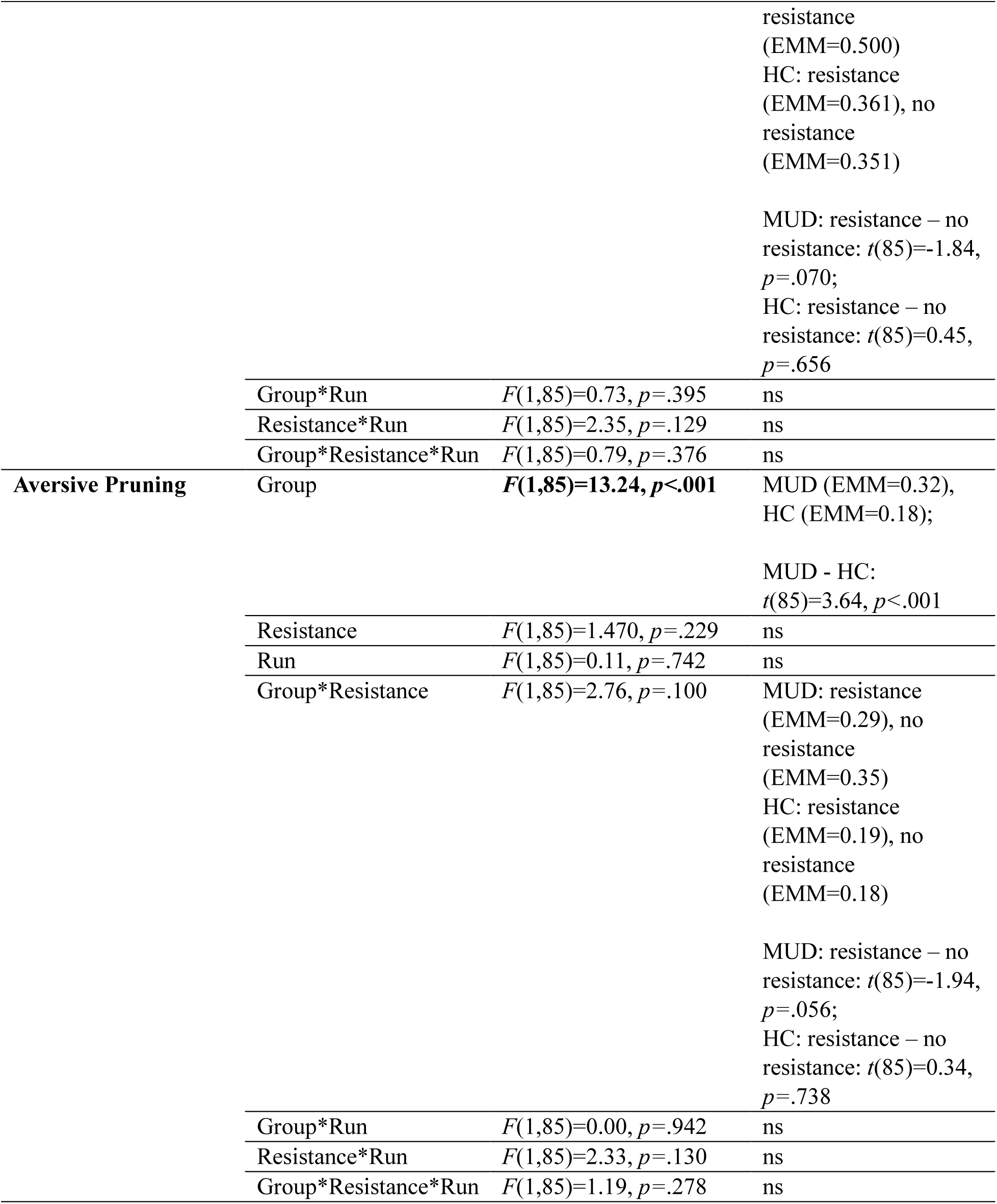
Full results of the models including run order for each computational parameter.

**Table S6.** Descriptive statistics and group comparison (where possible) for additional dimensional measures.

| Measure | HCs (*n*=49) | iMUDs (*n*=40) | Statistic |
| --- | --- | --- | --- |
| **CRT\*** | 3.27 (2.19) | 0.92 (1.29) | *t*(86)=5.92, *p*<.001, *d*=1.27 |
| **UPPS-P** |  |  |  |
| Negative Urgency | 22.16 (5.44) | 33.13 (6.25) | *t*(87)=8.84, *p*<.001, *d*=1.88 |
| Positive Urgency | 24.90 (5.11) | 34.08 (7.21) | *t*(87)=7.02, *p*<.001, *d*=1.50 |
| Lack of Perseverance | 18.49 (4.77) | 19.88 (6.08) | *t*(87)=1.20, *p*=.232, *d*=0.26 |
| Lack of Premeditation | 21.96 (4.37) | 22.60 (6.13) | *t*(87)=0.58, *p*=.567, *d*=0.12 |
| Sensation Seeking | 31.80 (6.53) | 36.68 (5.45) | *t*(87)=3.77, *p*<.001, *d*=0.80 |
| **BIS/BAS** |  |  |  |
| BIS | 18.74 (4.01) | 21.00 (3.40) | *t*(87)=2.83, *p*=.006, *d*=0.60 |
| Drive | 10.04 (2.00) | 12.20 (2.54) | *t*(87)=4.48, *p*<.001, *d*=0.96 |
| Reward-Seeking | 16.63 (1.52) | 17.58 (2.17) | *t*(87)=2.40, *p*=.018, *d*=0.51 |
| Fun-Seeking | 10.88 (2.22) | 12.65 (2.26) | *t*(87)=3.71, *p*<.001, *d*=0.79 |
| **TEPS** |  |  |  |
| Anticipatory | 45.71 (5.10) | 44.90 (6.55) | *t*(87)=0.66, *p*=.511, *d*=0.14 |
| Consummatory | 34.55 (4.21) | 34.25 (4.10) | *t*(87)=0.34, *p*=.735, *d*=0.07 |
| **DSQ†** | -- | 97.29 (34.58) | -- |
| **MAWQ** |  |  |  |
| Physical | -- | 0.98 (1.91) | -- |
| Emotional | -- | 2.13 (2.58) | -- |
| Functional | -- | 1.83 (2.31) | -- |
| Additional | -- | 2.73 (2.48) | -- |

\**n*=39 iMUDs; †*n*=28 iMUDs; *d*=Cohen's *d*

**Table S7.**
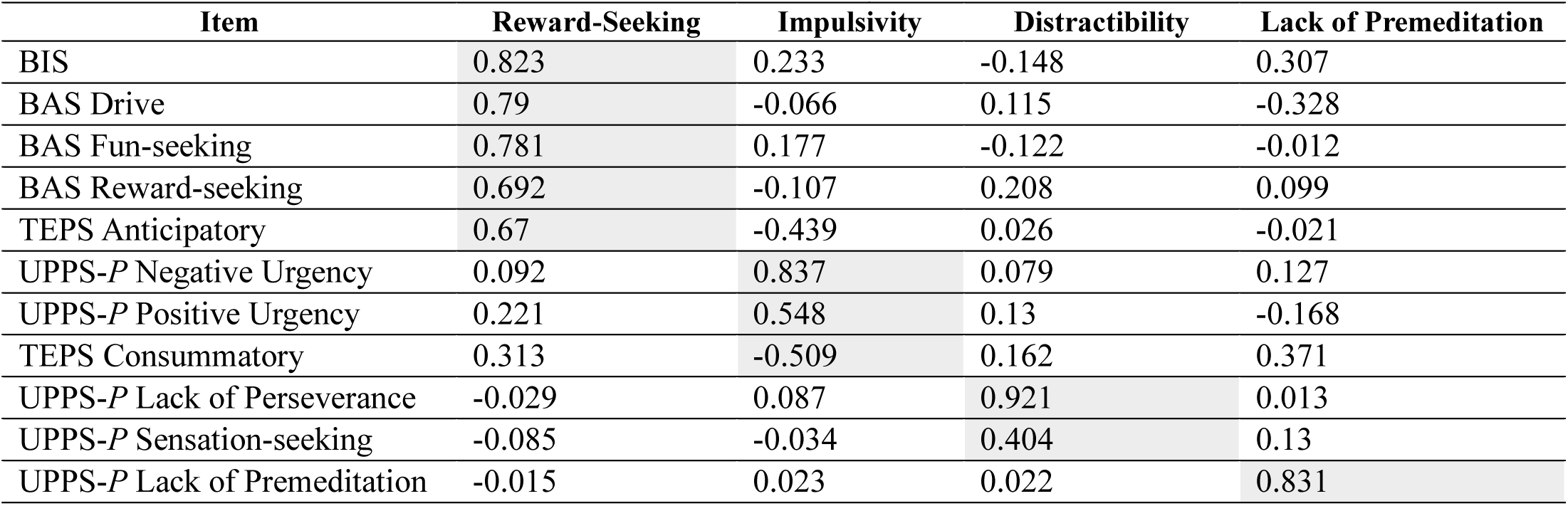
Factor scores for measures from latent factor analysis.

